# Accurate long-read sequencing identified GBA variants as a major genetic risk factor in the Luxembourg Parkinson’s study

**DOI:** 10.1101/2023.03.29.23287880

**Authors:** Sinthuja Pachchek Peiris, Zied Landoulsi, Lukas Pavelka, Claudia Schulte, Elena Buena-Atienza, Caspar Gross, Ann-Kathrin Hauser, Dheeraj Reddy Bobbili, Nicolas Casadei, Patrick May, Rejko Krüger, the NCER-PD Consortium

**Author notes:** **Corresponding authors:** Prof. Dr. Rejko Krüger, Luxembourg Centre for Systems Biomedicine (LCSB), University of Luxembourg, 6, avenue du Swing, L-4367 Belvaux, Luxembourg, Dr. Patrick May, Luxembourg Centre for Systems Biomedicine (LCSB), University of Luxembourg, 6, avenue du Swing, L-4367 Belvaux, Luxembourg.

## Abstract

Heterozygous variants in the glucocerebrosidase *GBA* gene are an increasingly recognized risk factor for Parkinson’s disease (PD). Due to the pseudogene *GBAP1* that shares 96% sequence homology with the *GBA* coding region, accurate variant calling by array-based or short-read sequencing methods remains a major challenge in understanding the genetic landscape of *GBA*-related PD. We established a novel long-read sequencing technology for assessing the full length of the *GBA* gene. We used subsequent regression models for genotype-phenotype analyses. We sequenced 752 patients with parkinsonism and 806 healthy controls of the Luxembourg Parkinson’s study. All *GBA* variants identified showed a 100% true positive rate by Sanger validation. We found 12% of unrelated PD patients carrying *GBA* variants. Three novel variants of unknown significance (VUS) were identified. Using a structure-based approach, we defined a potential risk prediction method for VUS. This study describes the full landscape of *GBA*-related parkinsonism in Luxembourg, showing a high prevalence of *GBA* variants as the major genetic risk for PD. Our approach provides an important advancement for highly accurate *GBA* variant calling, which is essential for providing access to emerging causative therapies for *GBA* carriers.

## 1. Introduction

Heterozygous variants in the glucocerebrosidase (*GBA*) gene, which encodes the enzyme β-glucocerebrosidase (GCase), are increasingly recognized as the most common genetic risk factor for the development of Parkinson’s disease (PD). Homozygous mutations in *GBA* are causative for the most frequent autosomal-recessive lysosomal storage disorder, Gaucher disease (GD).^1^ GD is characterized by a deficiency of the enzyme GCase which is necessary to hydrolyse the β-glucosyl linkage of glucosylceramide lipide (GlcCer) in lysosomes, to yield glucose and ceramide.^2^

The accurate variant calling in the *GBA* gene is challenging due to the presence of the highly homogeneous untranslated pseudogene called *GBAP1*, which is located 16 kilobases (kbp) downstream,^3^ and shares 96% sequence homology within the coding region.^4^ Furthermore, recombination and structural chromosomal variations within and around the *GBA* locus make the analysis more challenging.^5^ Complex alleles, which include several point mutations, are derived from a recombination between functional *GBA* and pseudogene *GBAP1*.^6^ RecNciI is the most prevalent recombinant allele, including the amino acid changes p.L483P and p.A495P, and the synonymous variant p.V499V.^6^

Our study aimed at accurately assessing all coding variants in the *GBA* gene among all participants of the Luxembourg Parkinson’s study,^7^ a case and control cohort including people with PD and atypical parkinsonism. To assess the accuracy of the novel targeted *GBA* sequencing method using Pacific Biosciences (PacBio)^8^ technology, we compared this method to genotyping with the NeuroChip array^9^ and short-read whole genome sequencing (WGS) data using Sanger sequencing as the gold standard for validation. We identified several types of pathogenic *GBA* variants (severe, mild, and risk) and further characterized genotype-phenotype associations to better understand the influence of each variant type and their effect on disease severity.

## 2. Results

### 2.1. Demographic and clinical characteristics

A total of 752 patients (652 PD patients and 100 patients with other forms of parkinsonism) and 806 HC were included (Figure 1). All participants were genotyped using NeuroChip and screened for *GBA* variants using targeted PacBio method, while a subset of 72 patients was screened with WGS. Among the patients, 66.4% (n = 499) were male with a mean age at disease onset of 63.1 ± 16 years. The control group consisted of 52.9% (n = 426) males with a mean age at assessment of 59.3 ± 12.2 years (Supplementary Table 1).

**Figure 1:**
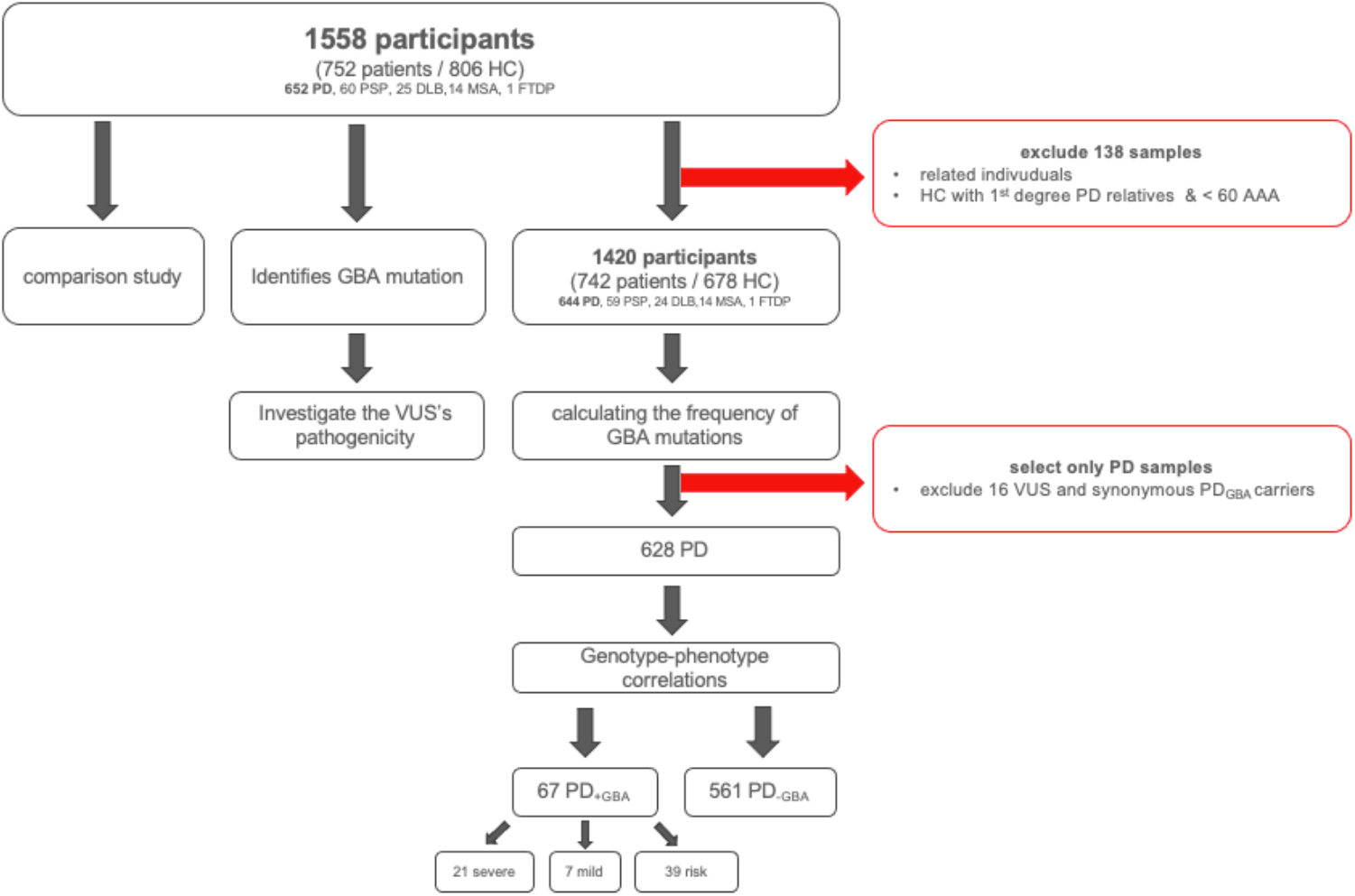
Description of the study dataset and methodology.

### 2.2. Targeted PacBio sequencing showed the highest specificity to detect *GBA* variants

To measure the reliability of *GBA* variant detection, we proceeded with two types of comparison. We compared PacBio, WGS and NeuroChip methods for a subset of samples (n=72). Then, we compared the PacBio and NeuroChip methods since they both covered most of the samples (n=1558). We considered as true positives only variants validated via Sanger sequencing (Supplementary Table 2).

First, we evaluated 72 samples screened by all three methods (Figure 2). Using the GBA-targeted PacBio method, we detected six individuals carrying *GBA* variants (p.E365K (n = 3), p.T408M (n = 1), p.N409S (n = 1), RecNciI, n = 1)). All the detected variants were confirmed by Sanger sequencing (true positive rate (TPR) of 100%). With the WGS method, we did not identify any false positive variant call, however, the WGS method failed to detect the RecNciI recombinant allele in one individual (TPR of 83.3% (5/6). Using Neurochip, we detected three potential *GBA* variants carriers (p.T408M (n = 1), p.N431S (n = 1), p.A215D (n=1), however, only one variant (p.T408M) was subsequently confirmed by Sanger sequencing (TPR of 16.6% (1/6) translating into a false detection rate (FDR) of 66.6% (2/3)). Next, we compared the results from 1558 samples screened with both, the *GBA* targeted PacBio method and the NeuroChip array (Figure 3). Using the GBA-targeted PacBio method, we detected 133 *GBA* variants carriers, of which 100% were validated by Sanger sequencing. Through the NeuroChip array, we detected 47 potential *GBA* variant carriers, among which only 36 were validated by Sanger sequencing (TPR of 27% (36/133), resulting in an FDR of 23.4% (11/47)).

**Figure 2.**
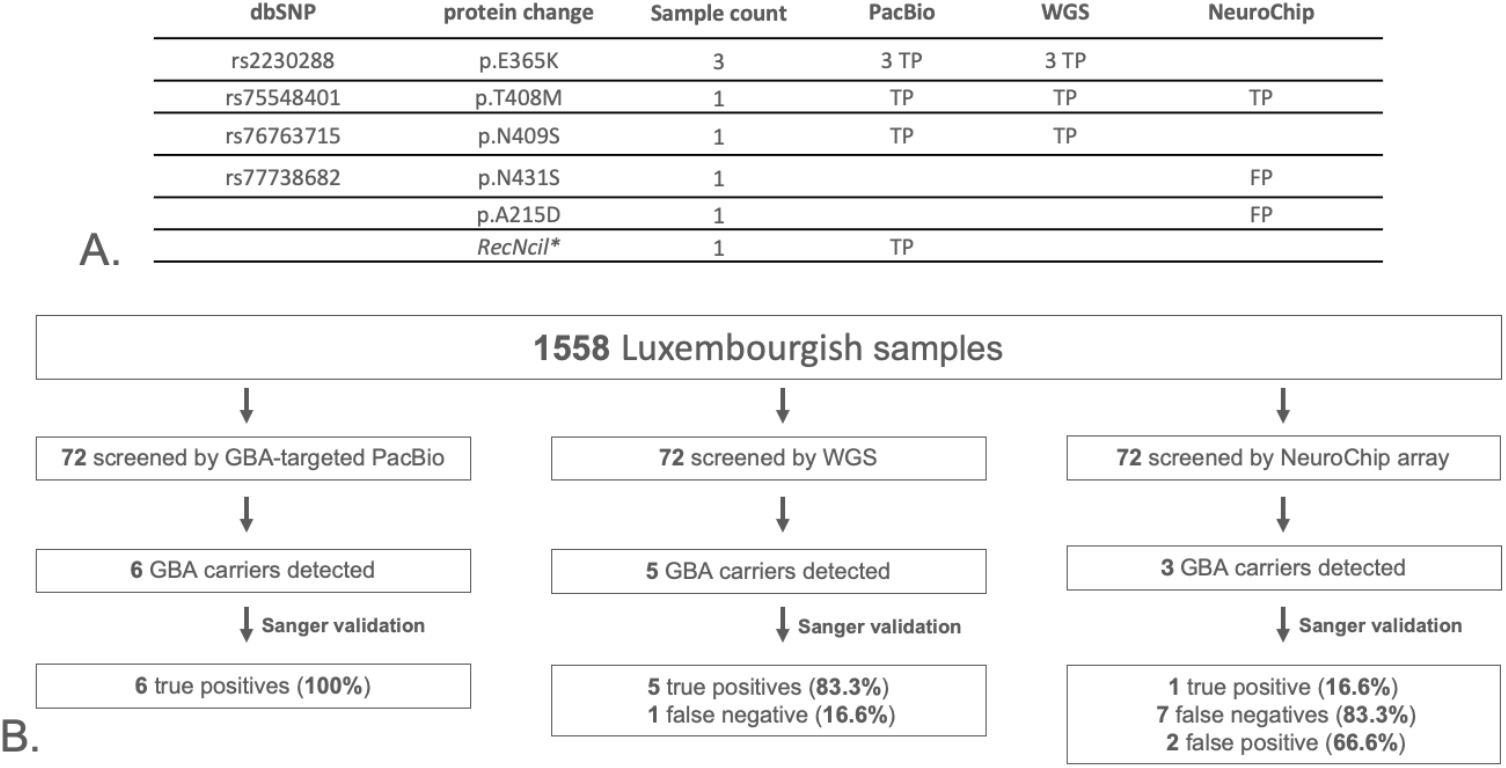
Comparison of variant calls from PacBio, WGS and NeuroChip genotyping data using 72 matched samples for the *GBA* gene and validated by Sanger sequencing. A) \**Rec*Nci*l (p*.*L483P; p*.*A495P; p*.*V499V)*; Sanger sequencing results : TP, true positive; FP, false positive. Sample count gives total number of samples carrying the variant found by each method. B) Comparative study of *GBA* variants detection by the GBA-targeted PacBio and NeuroChip array methods in the Luxembourg Parkinson’s study. Due to overrepresented variants with the NeuroChip array, we applied for the detected variants a study-wide threshold of 1% in our cohort.

**Figure 3:**
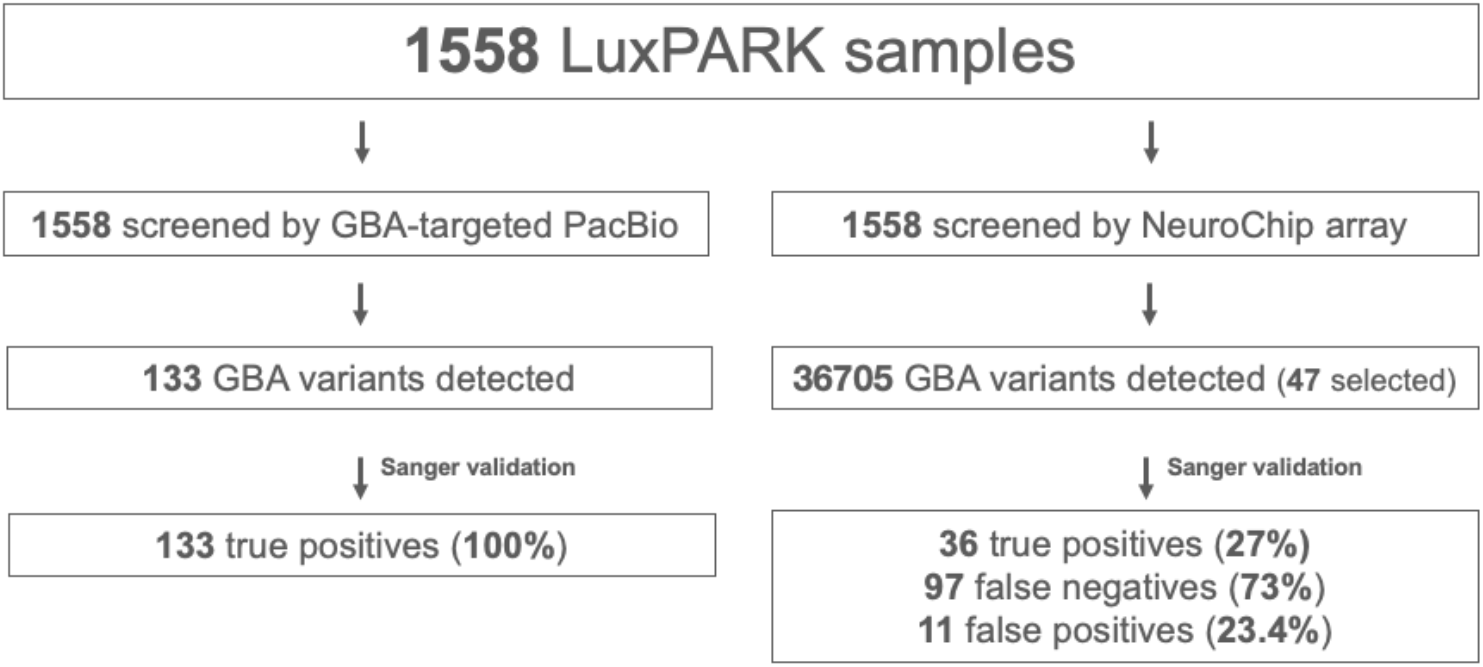
Comparative study of GBA variants detection by the GBA-targeted PacBio and NeuroChip array methods in the Luxembourg Parkinson’s study. Due to overrepresented variants with the NeuroChip array, we applied for the detected variants a study-wide threshold of 1% in our cohort.

### 2.3. Classification of *GBA* variants

From the 1558 individuals sequenced with the GBA-targeted PacBio method, we identified 124 carriers with at least one *GBA* variant (Supplementary Table 3). *GBA* variants were mostly heterozygous missense, one patient carried a heterozygous stop-gain variant p.R398*(rs121908309), two PD patients carried a homozygous missense variant p.E365K/p.E365K(rs2230288). We also detected nine different synonymous variants in exonic regions (Supplementary Table 4). The variant p.T408T(rs138498426) is a splice site variant (located within 2bp of the exon boundary) and classified as VUS.^10^ The remaining synonymous variants were not further analysed. Additionally, we identified 69 variants in intronic and UTRs regions (Supplementary Table 5) with unclear pathogenic relevance, of which 35 were rare. Based on Neurochip and WGS data, none of the *GBA* carriers carried pathogenic mutations in other PD associated genes as defined by MDSGene.^11^

We classified four combinations of multiple variants per individual as severe (p.N409S-p.L483P; the recombinant allele RecNciI; p.K13R-p.L483P; p.F252I-p.T408M) and one combination of variants as risk (Y61H-T408M) based on the classification of the respective associated pathogenic variants (Table 1).

**Table 1.**
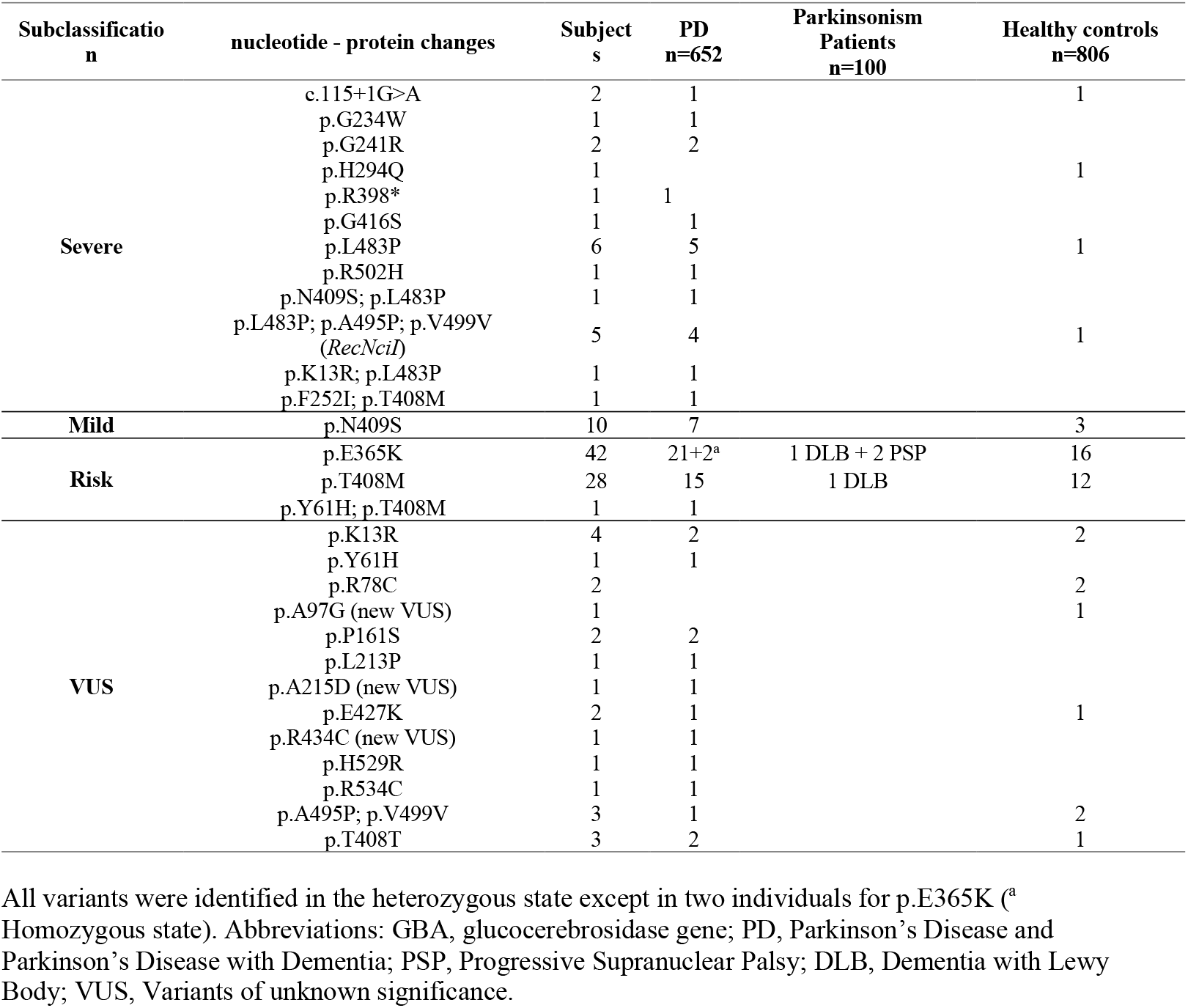
Distribution of *GBA* variants in the Luxembourg Parkinson’s study.

Overall, we detected 12% (77/644) *GBA* variant carriers among 644 unrelated PD patients and 5% (34/678) in healthy control individuals. We found a frequency of 10.4% (67/644) known pathogenic mutations in PD patients and 4.3% (29/678) in the control group (Table 2). Carriers of severe *GBA* mutations (n=21; OR=11.4; 95% CI=[2.6, 48.8]; *p*=0.0010) and risk *GBA* variants (n=39; OR=1.6; 95% CI=[1, 2.8]; *p*=0.0470) had a different risk of developing PD as defined by the indicated OR.

**Table 2.**
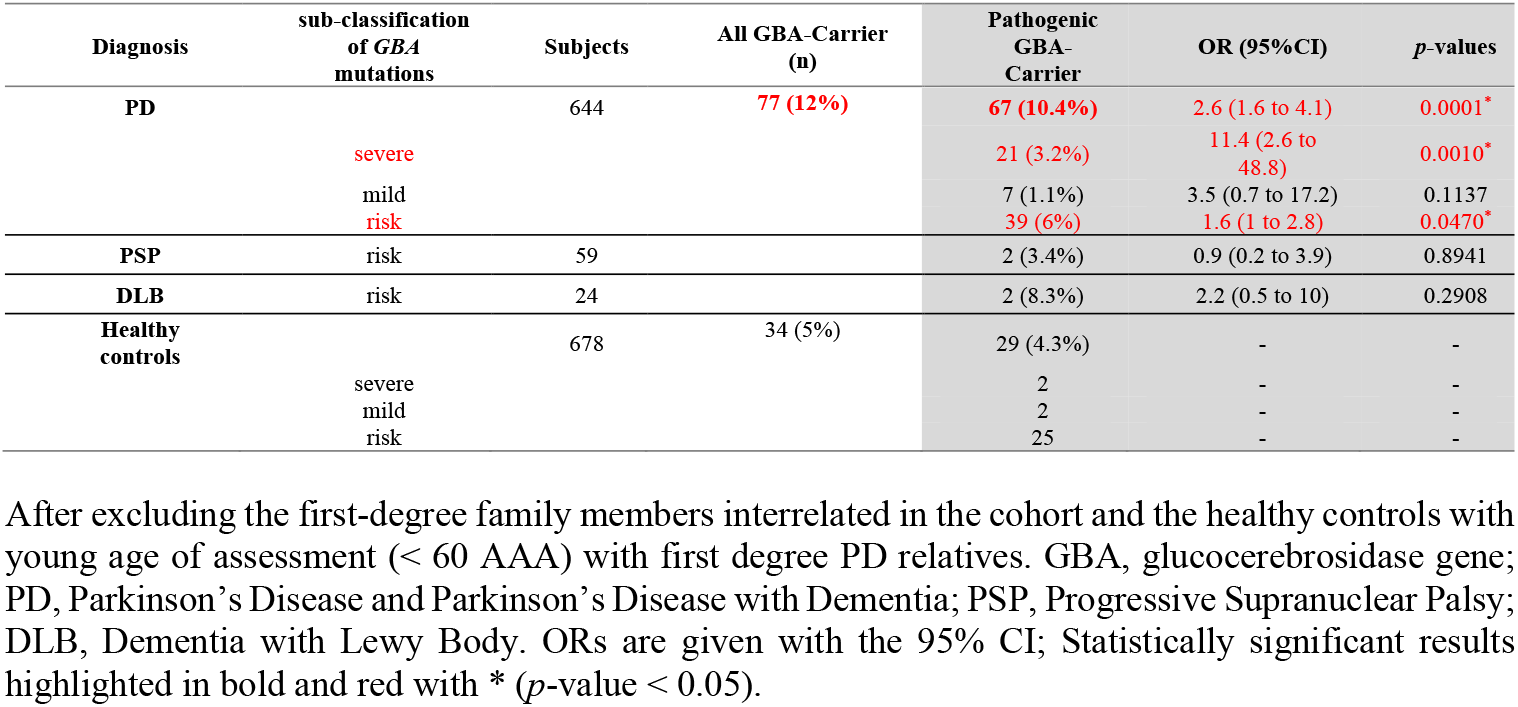
Frequency of GBA mutations in the Luxembourg Parkinson’s study.

| Diagnosis | sub-classification<br>of *GBA*<br>mutations | Subjects | All GBA-Carrier<br>(n) | Pathogenic<br>GBA-Carrier | OR (95%CI) | *p*-values |
| --- | --- | --- | --- | --- | --- | --- |
| PD |  | 644 | **77 (12%)** | **67 (10.4%)** | 2.6 (1.6 to 4.1) | 0.0001* |
|  | severe |  |  | 21 (3.2%) | 11.4 (2.6 to 48.8) | 0.0010* |
|  | mild |  |  | 7 (1.1%) | 3.5 (0.7 to 17.2) | 0.1137 |
|  | risk |  |  | 39 (6%) | 1.6 (1 to 2.8) | 0.0470* |
| PSP | risk | 59 |  | 2 (3.4%) | 0.9 (0.2 to 3.9) | 0.8941 |
| DLB | risk | 24 |  | 2 (8.3%) | 2.2 (0.5 to 10) | 0.2908 |
| Healthy<br>controls |  | 678 | 34 (5%) | 29 (4.3%) | - | - |
|  | severe |  |  | 2 | - | - |
|  | mild |  |  | 2 | - | - |
|  | risk |  |  | 25 | - | - |

After excluding the first-degree family members interrelated in the cohort and the healthy controls with young age of assessment (< 60 AAA) with first degree PD relatives. GBA, glucocerebrosidase gene; PD, Parkinson's Disease and Parkinson's Disease with Dementia; PSP, Progressive Supranuclear Palsy; DLB, Dementia with Lewy Body. ORs are given with the 95% CI; Statistically significant results highlighted in bold and red with \* (*p*-value < 0.05).

The most common *GBA* variants in PD patients were the risk variants p.E365K (n=23;3.5%) and p.T408M (n=17;2.6%).

### 2.4. Genotype-phenotype associations in GBA-PD patients

We characterized the clinical phenotype of severe, mild and risk *GBA* carriers and non-carriers only in unrelated PD patients excluding carriers with only one synonymous or VUS variants. The AAO was similar between *GBA* carriers (61.6±11.5) and non-carriers (62.5±11). Severe PD_GBA_ mutations carriers showed a trend towards younger AAO compared to mild and risk (severe: 58.6±13.1 vs mild: 65.4±17 vs risk: 62.5±0.3 years; *p*=0.29) (Table 3), with a significant risk to develop early onset PD (OR=3.76;*p*=0.0135).

**Table 3.** Demographic data for the PD patients in the Luxembourgish cohort separated by GBA mutation status.

| Features | All pathogenic variants<br>(n = 67) | Severe<br>(n = 21) | Mild<br>(n = 7) | Risk<br>(n = 39) | Noncarriers<br>(n = 561) |
| --- | --- | --- | --- | --- | --- |
| AAA, mean (SD) | 66.5 (±10.2)<br>[OR=0.37; p=0.4713] | 65.1 (±10.2)<br>[OR=0.1; p=0.3315] | 67.1 (±15.6)<br>[OR=0.68; p=0.9275] | 67.1 (±9.2)<br>[OR=0.67; p=0.8231] | 67.5 (±10.9) |
| Sex, Male % (n) | 40 (59.7%)<br>[OR=0.71; p=0.1882] | 13 (61.9%)<br>[OR=0.77; p=0.5762] | 5 (71.4%)<br>[OR=1.19; p=0.8356] | 22 (56.4%)<br>[OR=0.62; p=0.149] | 380 (67.7%) |
| AAO, mean (SD) | 61.6 (±11.5)<br>[OR=0.42; p=0.5668] | 58.6 (±13.1)<br>[OR=0.02; p=0.1383] | 65.4 (±17.0)<br>[OR=19.33; p=0.511] | 62.5 (±9.3)<br>[OR=1.07; p=0.9704] | 62.5 (±11.8) |
| AAO ≤ 45 | 8 (11.9%)<br>[OR=1.63; p=0.2301] | 5 (23.8%)<br>[OR=3.76; p=0.0135*] | 2 (28.6%)<br>[OR=4.82; p=0.0648] | 1 (2.6%)<br>[OR=0.32; p=0.2626] | 43 (7.7%) |
| Disease Duration, mean (SD) | 4.7 (±4.8)<br>[OR=0.7; p=0.7074] | 6.4 (±4.7)<br>[OR=3.99; p=0.2338] | 1.7 (±1.4)<br>[OR=0.04; p=0.0979] | 4.4 (±4.9)<br>[OR=0.56; p=0.4966] | 5 (±5.2) |
| Family History, N (%) | 25 (37.3%)<br>[OR=1.5; p=0.1137] | 8 (38.1%)<br>[OR=1.58; p=0.3167] | 2 (28.6%)<br>[OR=1.03; p=0.9726] | 15 (38.5%)<br>[OR=1.61; p=0.1651] | 157 (28%) |

After excluding the first-degree family members interrelated in the cohort, the healthy controls with young age of assessment (< 60 AAA) with first degree PD relatives and synonymous and VUS variants carriers. Data are given as mean (SD) or N (%). Significance level for comparison is $p < 0.05$ . AAA, age at assessment in years ; AAO, Age at onset in years.

We compared clinical features between PD patients carrying pathogenic *GBA* variants and PD patients without *GBA* variants. We found that the sense of smell was strongly impaired in carriers (uncorrected *p*=0.0198) (Supplementary Table 6). Next, we compared patients carrying variants from each category (severe, mild or risk) separately with PD patients without *GBA* variants (Table 4). Carriers of severe *GBA* mutation showed more severe non-motor symptoms when compared to non-GBA carriers, such as MDS-UPDRS Part I (uncorrected *p*=0.0088) and hallucinations (uncorrected *p*=0.015), and also an impaired sense of smell as assessed by Sniffin’ Stick test (uncorrected *p*=0.0403). To show the deleterious impact of the severe variants, we compared carriers of severe variants with patients carrying either mild or risk *GBA* variants (Table 5). We observed here that severe variants carriers have more severe gait disorder and depression and worse MDS-UPDRS Part I and PDQ-39. For all clinical features, there were no significant associations after the correction for multiple comparisons using FDR adjustment.

**Table 4.**
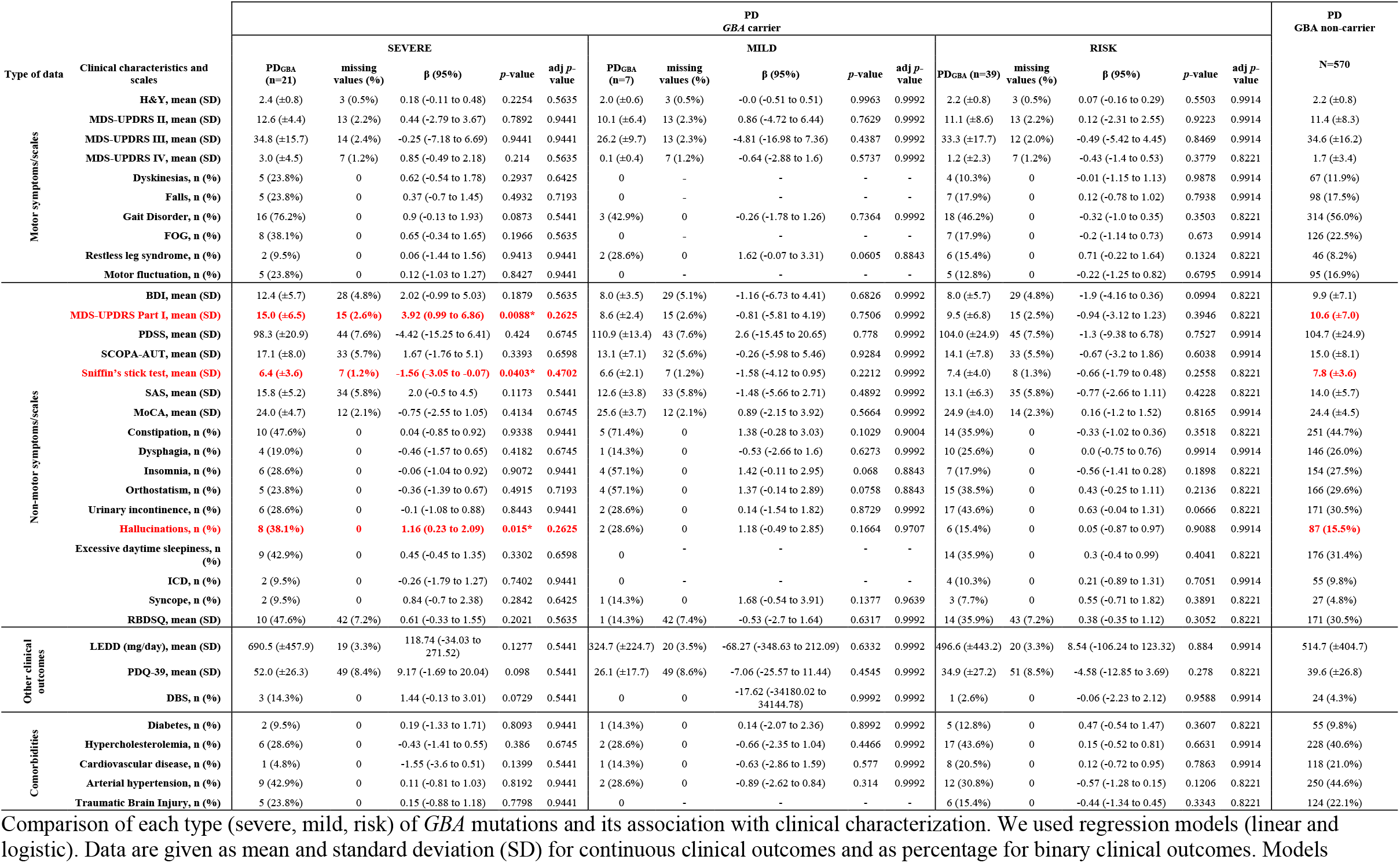
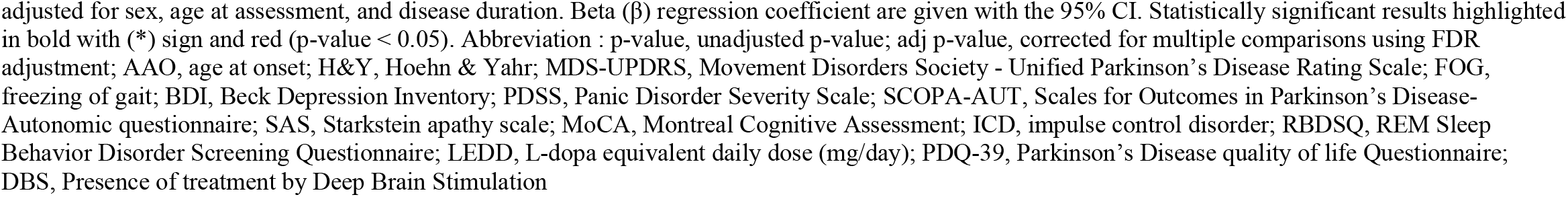
Clinical characteristics of PD classified by GBA mutation status.

| Type of data | Clinical characteristics and scales | PD<br>GBA carrier |  |  |  |  |  |  |  |  |  |  |  |  |  | PD<br>GBA non-carrier |  |
| --- | --- | --- | --- | --- | --- | --- | --- | --- | --- | --- | --- | --- | --- | --- | --- | --- | --- |
|  |  | SEVERE |  |  |  |  | MILD |  |  |  |  | RISK |  |  |  |  | N=570 |
|  |  | PD<sub>GBA</sub> (n=21) | missing values (%) | β (95%) | p-value | adj p-value | PD<sub>GBA</sub> (n=7) | missing values (%) | β (95%) | p-value | adj p-value | PD<sub>GBA</sub> (n=39) | missing values (%) | β (95%) | p-value | adj p-value |  |
| Motor symptoms/scales | H&Y, mean (SD) | 2.4 (±0.8) | 3 (0.5%) | 0.18 (-0.11 to 0.48) | 0.2254 | 0.5635 | 2.0 (±0.6) | 3 (0.5%) | -0.0 (-0.51 to 0.51) | 0.9963 | 0.9992 | 2.2 (±0.8) | 3 (0.5%) | 0.07 (-0.16 to 0.29) | 0.5503 | 0.9914 | 2.2 (±0.8) |
|  | MDS-UPDRS II, mean (SD) | 12.6 (±4.4) | 13 (2.2%) | 0.44 (-2.79 to 3.67) | 0.7892 | 0.9441 | 10.1 (±6.4) | 13 (2.3%) | 0.86 (-4.72 to 6.44) | 0.7629 | 0.9992 | 11.1 (±8.6) | 13 (2.2%) | 0.12 (-2.31 to 2.55) | 0.9223 | 0.9914 | 11.4 (±8.3) |
|  | MDS-UPDRS III, mean (SD) | 34.8 (±15.7) | 14 (2.4%) | -0.25 (-7.18 to 6.69) | 0.9441 | 0.9441 | 26.2 (±9.7) | 13 (2.3%) | -4.81 (-16.98 to 7.36) | 0.4387 | 0.9992 | 33.3 (±17.7) | 12 (2.0%) | -0.49 (-5.42 to 4.45) | 0.8469 | 0.9914 | 34.6 (±16.2) |
|  | MDS-UPDRS IV, mean (SD) | 3.0 (±4.5) | 7 (1.2%) | 0.85 (-0.49 to 2.18) | 0.214 | 0.5635 | 0.1 (±0.4) | 7 (1.2%) | -0.64 (-2.88 to 1.6) | 0.5737 | 0.9992 | 1.2 (±2.3) | 7 (1.2%) | -0.43 (-1.4 to 0.53) | 0.3779 | 0.8221 | 1.7 (±3.4) |
|  | Dyskinesias, n (%) | 5 (23.8%) | 0 | 0.62 (-0.54 to 1.78) | 0.2937 | 0.6425 | 0 | - | - | - | - | 4 (10.3%) | 0 | -0.01 (-1.15 to 1.13) | 0.9878 | 0.9914 | 67 (11.9%) |
|  | Falls, n (%) | 5 (23.8%) | 0 | 0.37 (-0.7 to 1.45) | 0.4932 | 0.7193 | 0 | - | - | - | - | 7 (17.9%) | 0 | 0.12 (-0.78 to 1.02) | 0.7938 | 0.9914 | 98 (17.5%) |
|  | Gait Disorder, n (%) | 16 (76.2%) | 0 | 0.9 (-0.13 to 1.93) | 0.0873 | 0.5441 | 3 (42.9%) | 0 | -0.26 (-1.78 to 1.26) | 0.7364 | 0.9992 | 18 (46.2%) | 0 | -0.32 (-1.0 to 0.35) | 0.3503 | 0.8221 | 314 (56.0%) |
|  | FOG, n (%) | 8 (38.1%) | 0 | 0.65 (-0.34 to 1.65) | 0.1966 | 0.5635 | 0 | - | - | - | - | 7 (17.9%) | 0 | -0.2 (-1.14 to 0.73) | 0.673 | 0.9914 | 126 (22.5%) |
|  | Restless leg syndrome, n (%) | 2 (9.5%) | 0 | 0.06 (-1.44 to 1.56) | 0.9413 | 0.9441 | 2 (28.6%) | 0 | 1.62 (-0.07 to 3.31) | 0.0605 | 0.8843 | 6 (15.4%) | 0 | 0.71 (-0.22 to 1.64) | 0.1324 | 0.8221 | 46 (8.2%) |
|  | Motor fluctuation, n (%) | 5 (23.8%) | 0 | 0.12 (-1.03 to 1.27) | 0.8427 | 0.9441 | 0 | - | - | - | - | 5 (12.8%) | 0 | -0.22 (-1.25 to 0.82) | 0.6795 | 0.9914 | 95 (16.9%) |
| Non-motor symptoms/scales | BDI, mean (SD) | 12.4 (±5.7) | 28 (4.8%) | 2.02 (-0.99 to 5.03) | 0.1879 | 0.5635 | 8.0 (±3.5) | 29 (5.1%) | -1.16 (-6.73 to 4.41) | 0.6826 | 0.9992 | 8.0 (±5.7) | 29 (4.8%) | -1.9 (-4.16 to 0.36) | 0.0994 | 0.8221 | 9.9 (±7.1) |
|  | MDS-UPDRS Part I, mean (SD) | 15.0 (±6.5) | 15 (2.6%) | 3.92 (0.99 to 6.86) | 0.0088* | 0.2625 | 8.6 (±2.4) | 15 (2.6%) | -0.81 (-5.81 to 4.19) | 0.7506 | 0.9992 | 9.5 (±6.8) | 15 (2.5%) | -0.94 (-3.12 to 1.23) | 0.3946 | 0.8221 | 10.6 (±7.0) |
|  | PDSS, mean (SD) | 98.3 (±20.9) | 44 (7.6%) | -4.42 (-15.25 to 6.41) | 0.424 | 0.6745 | 110.9 (±13.4) | 43 (7.6%) | 2.6 (-15.45 to 20.65) | 0.778 | 0.9992 | 104.0 (±24.9) | 45 (7.5%) | -1.3 (-9.38 to 6.78) | 0.7527 | 0.9914 | 104.7 (±24.9) |
|  | SCOPA-AUT, mean (SD) | 17.1 (±8.0) | 33 (5.7%) | 1.67 (-1.76 to 5.1) | 0.3393 | 0.6598 | 13.1 (±7.1) | 32 (5.6%) | -0.26 (-5.98 to 5.46) | 0.9284 | 0.9992 | 14.1 (±7.8) | 33 (5.5%) | -0.67 (-3.2 to 1.86) | 0.6038 | 0.9914 | 15.0 (±8.1) |
|  | Sniffin's stick test, mean (SD) | 6.4 (±3.6) | 7 (1.2%) | -1.56 (-3.05 to -0.07) | 0.0403* | 0.4702 | 6.6 (±2.1) | 7 (1.2%) | -1.58 (-4.12 to 0.95) | 0.2212 | 0.9992 | 7.4 (±4.0) | 8 (1.3%) | -0.66 (-1.79 to 0.48) | 0.2558 | 0.8221 | 7.8 (±3.6) |
|  | SAS, mean (SD) | 15.8 (±5.2) | 34 (5.8%) | 2.0 (-0.5 to 4.5) | 0.1173 | 0.5441 | 12.6 (±3.8) | 33 (5.8%) | -1.48 (-5.66 to 2.71) | 0.4892 | 0.9992 | 13.1 (±6.3) | 35 (5.8%) | -0.77 (-2.66 to 1.11) | 0.4228 | 0.8221 | 14.0 (±5.7) |
|  | MoCA, mean (SD) | 24.0 (±4.7) | 12 (2.1%) | -0.75 (-2.55 to 1.05) | 0.4134 | 0.6745 | 25.6 (±3.7) | 12 (2.1%) | 0.89 (-2.15 to 3.92) | 0.5664 | 0.9992 | 24.9 (±4.0) | 14 (2.3%) | 0.16 (-1.2 to 1.52) | 0.8165 | 0.9914 | 24.4 (±4.5) |
|  | Constipation, n (%) | 10 (47.6%) | 0 | 0.04 (-0.85 to 0.92) | 0.9338 | 0.9441 | 5 (71.4%) | 0 | 1.38 (-0.28 to 3.03) | 0.1029 | 0.9004 | 14 (35.9%) | 0 | -0.33 (-1.02 to 0.36) | 0.3518 | 0.8221 | 251 (44.7%) |
|  | Dysphagia, n (%) | 4 (19.0%) | 0 | -0.46 (-1.57 to 0.65) | 0.4182 | 0.6745 | 1 (14.3%) | 0 | -0.53 (-2.66 to 1.6) | 0.6273 | 0.9992 | 10 (25.6%) | 0 | 0.0 (-0.75 to 0.76) | 0.9914 | 0.9914 | 146 (26.0%) |
|  | Insomnia, n (%) | 6 (28.6%) | 0 | -0.06 (-1.04 to 0.92) | 0.9072 | 0.9441 | 4 (57.1%) | 0 | 1.42 (-0.11 to 2.95) | 0.068 | 0.8843 | 7 (17.9%) | 0 | -0.56 (-1.41 to 0.28) | 0.1898 | 0.8221 | 154 (27.5%) |
|  | Orthostatism, n (%) | 5 (23.8%) | 0 | -0.36 (-1.39 to 0.67) | 0.4915 | 0.7193 | 4 (57.1%) | 0 | 1.37 (-0.14 to 2.89) | 0.0758 | 0.8843 | 15 (38.5%) | 0 | 0.43 (-0.25 to 1.11) | 0.2136 | 0.8221 | 166 (29.6%) |
|  | Urinary incontinence, n (%) | 6 (28.6%) | 0 | -0.1 (-1.08 to 0.88) | 0.8443 | 0.9441 | 2 (28.6%) | 0 | 0.14 (-1.54 to 1.82) | 0.8729 | 0.9992 | 17 (43.6%) | 0 | 0.63 (-0.04 to 1.31) | 0.0666 | 0.8221 | 171 (30.5%) |
|  | Hallucinations, n (%) | 8 (38.1%) | 0 | 1.16 (0.23 to 2.09) | 0.015* | 0.2625 | 2 (28.6%) | 0 | 1.18 (-0.49 to 2.85) | 0.1664 | 0.9707 | 6 (15.4%) | 0 | 0.05 (-0.87 to 0.97) | 0.9088 | 0.9914 | 87 (15.5%) |
|  | Excessive daytime sleepiness, n (%) | 9 (42.9%) | 0 | 0.45 (-0.45 to 1.35) | 0.3302 | 0.6598 | 0 | - | - | - | - | 14 (35.9%) | 0 | 0.3 (-0.4 to 0.99) | 0.4041 | 0.8221 | 176 (31.4%) |
|  | ICD, n (%) | 2 (9.5%) | 0 | -0.26 (-1.79 to 1.27) | 0.7402 | 0.9441 | 0 | - | - | - | - | 4 (10.3%) | 0 | 0.21 (-0.89 to 1.31) | 0.7051 | 0.9914 | 55 (9.8%) |
|  | Syncope, n (%) | 2 (9.5%) | 0 | 0.84 (-0.7 to 2.38) | 0.2842 | 0.6425 | 1 (14.3%) | 0 | 1.68 (-0.54 to 3.91) | 0.1377 | 0.9639 | 3 (7.7%) | 0 | 0.55 (-0.71 to 1.82) | 0.3891 | 0.8221 | 27 (4.8%) |
|  | RBDSQ, mean (SD) | 10 (47.6%) | 42 (7.2%) | 0.61 (-0.33 to 1.55) | 0.2021 | 0.5635 | 1 (14.3%) | 42 (7.4%) | -0.53 (-2.7 to 1.64) | 0.6317 | 0.9992 | 14 (35.9%) | 43 (7.2%) | 0.38 (-0.35 to 1.12) | 0.3052 | 0.8221 | 171 (30.5%) |
| Other clinical outcomes | LEDD (mg/day), mean (SD) | 690.5 (±457.9) | 19 (3.3%) | 118.74 (-34.03 to 271.52) | 0.1277 | 0.5441 | 324.7 (±224.7) | 20 (3.5%) | -68.27 (-348.63 to 212.09) | 0.6332 | 0.9992 | 496.6 (±443.2) | 20 (3.3%) | 8.54 (-106.24 to 123.32) | 0.884 | 0.9914 | 514.7 (±404.7) |
|  | PDQ-39, mean (SD) | 52.0 (±26.3) | 49 (8.4%) | 9.17 (-1.69 to 20.04) | 0.098 | 0.5441 | 26.1 (±17.7) | 49 (8.6%) | -7.06 (-25.57 to 11.44) | 0.4545 | 0.9992 | 34.9 (±27.2) | 51 (8.5%) | -4.58 (-12.85 to 3.69) | 0.278 | 0.8221 | 39.6 (±26.8) |
|  | DBS, n (%) | 3 (14.3%) | 0 | 1.44 (-0.13 to 3.01) | 0.0729 | 0.5441 | 0 | 0 | -17.62 (-34180.02 to 34144.78) | 0.9992 | 0.9992 | 1 (2.6%) | 0 | -0.06 (-2.23 to 2.12) | 0.9588 | 0.9914 | 24 (4.3%) |
| Comorbidities | Diabetes, n (%) | 2 (9.5%) | 0 | 0.19 (-1.33 to 1.71) | 0.8093 | 0.9441 | 1 (14.3%) | 0 | 0.14 (-2.07 to 2.36) | 0.8992 | 0.9992 | 5 (12.8%) | 0 | 0.47 (-0.54 to 1.47) | 0.3607 | 0.8221 | 55 (9.8%) |
|  | Hypercholesterolemia, n (%) | 6 (28.6%) | 0 | -0.43 (-1.41 to 0.55) | 0.386 | 0.6745 | 2 (28.6%) | 0 | -0.66 (-2.35 to 1.04) | 0.4466 | 0.9992 | 17 (43.6%) | 0 | 0.15 (-0.52 to 0.81) | 0.6631 | 0.9914 | 228 (40.6%) |
|  | Cardiovascular disease, n (%) | 1 (4.8%) | 0 | -1.55 (-3.6 to 0.51) | 0.1399 | 0.5441 | 1 (14.3%) | 0 | -0.63 (-2.86 to 1.59) | 0.577 | 0.9992 | 8 (20.5%) | 0 | 0.12 (-0.72 to 0.95) | 0.7863 | 0.9914 | 118 (21.0%) |
|  | Arterial hypertension, n (%) | 9 (42.9%) | 0 | 0.11 (-0.81 to 1.03) | 0.8192 | 0.9441 | 2 (28.6%) | 0 | -0.89 (-2.62 to 0.84) | 0.314 | 0.9992 | 12 (30.8%) | 0 | -0.57 (-1.28 to 0.15) | 0.1206 | 0.8221 | 250 (44.6%) |
|  | Traumatic Brain Injury, n (%) | 5 (23.8%) | 0 | 0.15 (-0.88 to 1.18) | 0.7798 | 0.9441 | 0 | - | - | - | - | 6 (15.4%) | 0 | -0.44 (-1.34 to 0.45) | 0.3343 | 0.8221 | 124 (22.1%) |

Comparison of each type (severe, mild, risk) of *GBA* mutations and its association with clinical characterization. We used regression models (linear and logistic). Data are given as mean and standard deviation (SD) for continuous clinical outcomes and as percentage for binary clinical outcomes. Models
adjusted for sex, age at assessment, and disease duration. Beta ( $\beta$ ) regression coefficient are given with the 95% CI. Statistically significant results highlighted in bold with (\*) sign and red (p-value < 0.05). Abbreviation : p-value, unadjusted p-value; adj p-value, corrected for multiple comparisons using FDR adjustment; AAO, age at onset; H&Y, Hoehn & Yahr; MDS-UPDRS, Movement Disorders Society - Unified Parkinson's Disease Rating Scale; FOG, freezing of gait; BDI, Beck Depression Inventory; PDSS, Panic Disorder Severity Scale; SCOPA-AUT, Scales for Outcomes in Parkinson's Disease- Autonomic questionnaire; SAS, Starkstein apathy scale; MoCA, Montreal Cognitive Assessment; ICD, impulse control disorder; RBDSQ, REM Sleep Behavior Disorder Screening Questionnaire; LEDD, L-dopa equivalent daily dose (mg/day); PDQ-39, Parkinson's Disease quality of life Questionnaire; DBS, Presence of treatment by Deep Brain Stimulation

**Table 5.**
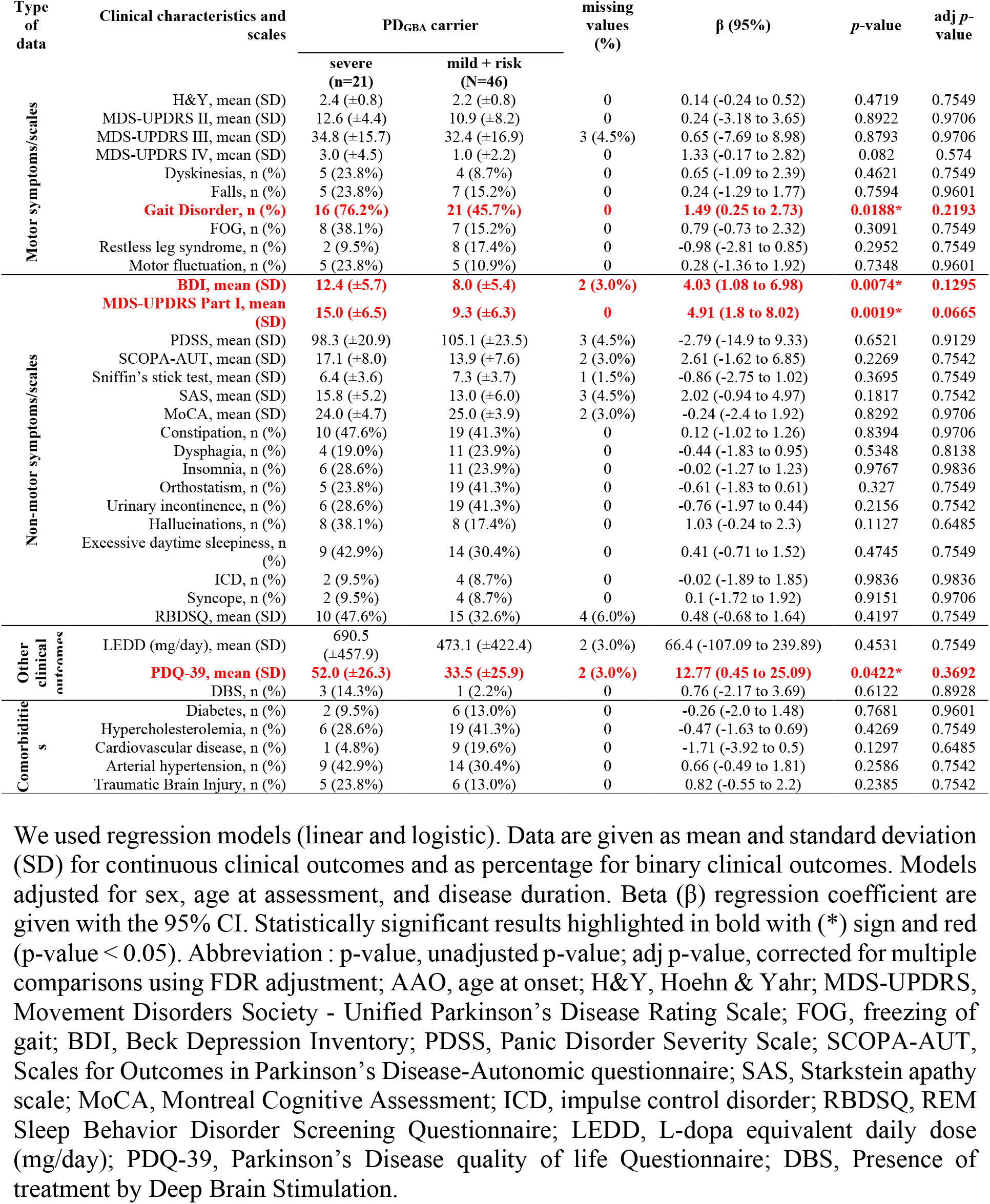
The deleterious impact of severe GBA-PD carriers in comparison with mild and risk and their clinical characteristics.

### 2.5. VUS and the Glucosylceramidase structure

We detected nine already reported VUS (p.K13R, p.Y61H, p.R78C, p.L213P, p.E427K, p.A495P, p.H529R, p.R534C, p.T408T) and three new VUS (p.A97G, p.A215 and p.R434C). According to our strategy developed for VUS *GBA* variants classification, where we assign the pathogenicity based on the REVEL, the CADD, the dbscSNV scores, as well as whether the patients carrying the variants. We suggest to sub-classify the variants p.Y61H, p.L213P, p.A215D, and p.R434C as severe variants. The variant p.L213P changes the Leucine amino acid into proline, which is known to be the ‘helix breaker’ amino acid that can induce a bend into the protein structure^12^(Supplementary Figure 1). The p.L213P and p.A215D variants are in the catalytic site of the enzyme in the triose-phosphate isomerase (TIM) barrel structure. The p.Y61H variant (Figure 4.A) is next in sequence and in structure to the known severe PD variant p.C62W and the patient carrying this variant had an AAO of 38 years, indicating an early-onset likely severe form of PD. The p. R434C variant is close to a known severe (p.V433L) and mild (p.W432R, p.N435T) PD variants in the 3D structure. We propose to sub-classify the variants p.H529R and p.R534C as mild, as they are both found only in PD patients. The variants p.K13R, p.R78C, p.E427K, and p.A495P are sub-classified as risk variants. The variant p.K13R is located in the signal peptide region. The variant p.R78C was annotated as “PD susceptibility” in HGMD with deleterious impact in CADD. The variant p.E427K was annotated as linked to “parkinsonism” in ClinVar and “reduced activity” in HGMD. We suggest to classify the variant p.A97G as probably benign because it is localized in a coil-bend structure and is not close to any known pathogenic variants.

**Figure 4.**
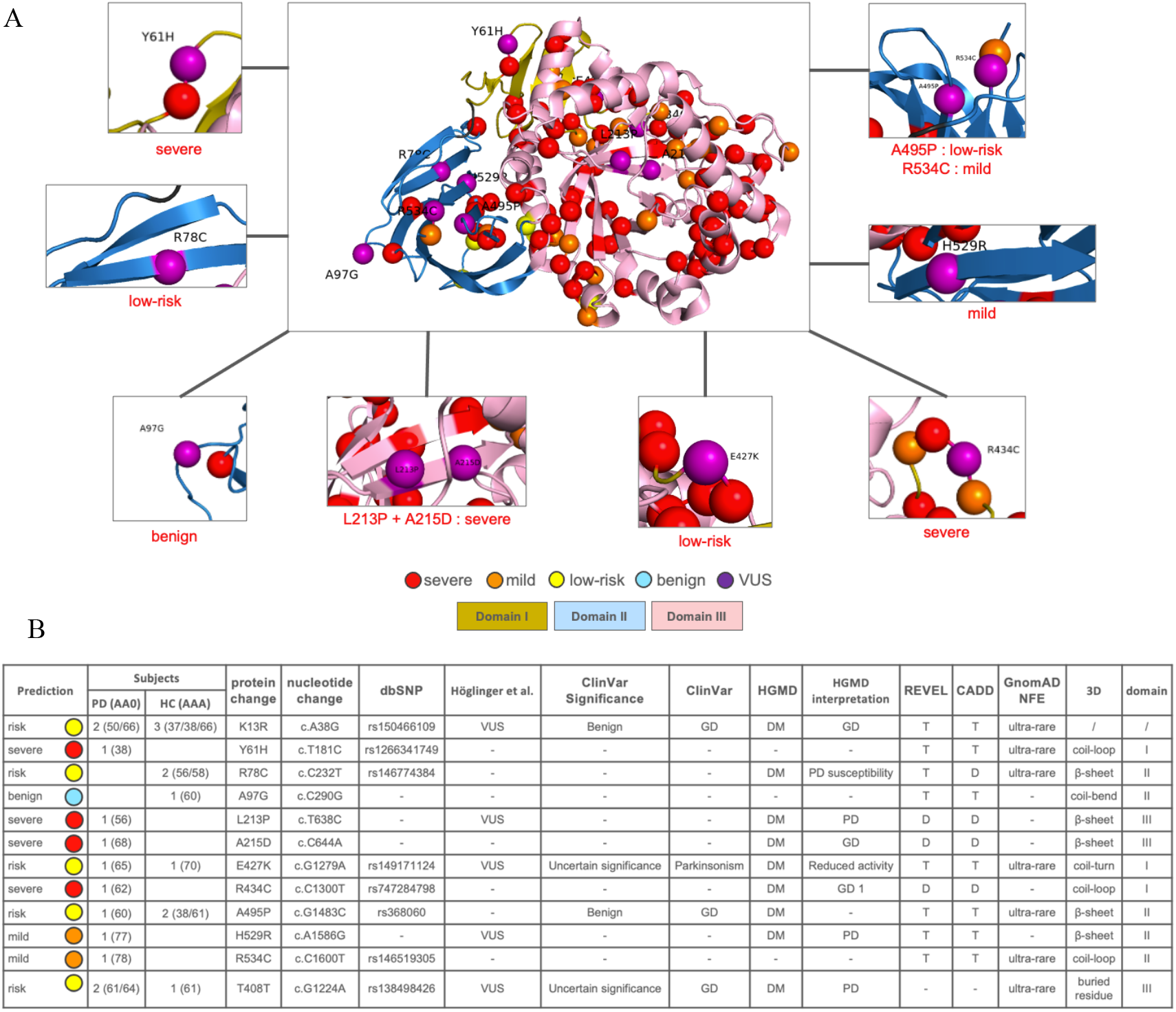
Sub-classification of VUS found in the Luxembourg Parkinson’s study. A) *GBA* missense and stop gain variants mapped onto the three-dimensional structure of GCase. Domain 1 is shown in dark yellow, domain 2 in blue, and domain 3 in pink. Variants classified as severe are coloured red, mild are coloured orange, risk in yellow and VUS are coloured purple. B) GBA, glucocerebrosidase gene; GD, Gaucher’s disease; PD, Parkinson’s disease. HGMD, The Human Gene Mutation Database; REVEL, Rare Exome Variant Ensemble Learner; CADD, Combined Annotation Dependent Depletion; gnomAD, The Genome Aggregation Database. DM, Disease causing mutation; D, Deleterious; T, Tolerate. Variants classified as severe are coloured red, mild are coloured orange, risk in yellow and VUS are coloured purple.

The synonymous variant p.T408T was found in two cases and one healthy control individual. Two established splice-site prediction scores (dbscSNV: ada_score 0.9797 and rf_score 0.85) agreed in their prediction that the variant is likely to affect splicing. HGMD classified the variant as disease mutation (DM) (Supplementary Table 4). Therefore, we propose to classify the variant as a risk variant.

In total, we propose to classify four VUS variants as severe (p.Y61H, p.L213P, p.A215D, and p.R434C), two as mild (p.H529R and p.R534C), five as risk (p.K13R, p.R78C, p.E427K, p.A495P and p.T408T) and one as benign (p.A97G) (Figure 4.B).

## 3. Discussion

Our study showed, for the first time, the utility of targeted PacBio sequencing as a highly sensitive and specific method to identify known and novel *GBA* variants. The PacBio method demonstrated a very high efficiency by targeting the entire length of the *GBA* gene with 100% reliability and solves the problems arising from the presence of the *GBAP1* pseudogene. The effectiveness of the target PacBio method to investigate relevant genes with homologous pseudogenes has also been proven in several other studies.^12–15^ The comparative study that we conducted with the different screening methods for *GBA* mutations will help researchers to be more accurate and comprehensive implying a more critical appraisal of the results obtained by NeuroChip and WGS with more false positive and false negative results.

*GBA* mutations were identified as the most common genetic risk factor for the development of PD. A heterozygous *GBA* variant was typically observed in 4%–12% of PD patients in different populations worldwide, with the highest prevalence of 20% described in Ashkenazi Jewish PD patients. ^16,17^ Important variation is due to ethnicity, the investigated mutations and the sequencing method used. Our study describes the landscape of *GBA* carriers in the studied Luxembourgish population showing the high prevalence of *GBA* mutations that could be the major genetic risk factor of PD in Luxembourg. The frequency of GBA mutation in PD in our study was 12% and we observed a significantly higher proportion of pathogenic (severe, mild and risk) *GBA* variants in PD patients compared to HC (10.4% *vs* 4.3%;OR=2.6;CI=[1.6,4.1],*p*=0.0001). Compared to previous studies, our study highlights that using the new PacBio sequencing method, the Luxembourg Parkinson’s study cohort showed a comparable frequency of PD_GBA_ carriers reported so far in similarly sized Italian^18^ and Spanish^19^ cohorts (Supplementary Table 7). When comparing previous reports of *GBA* variants in different populations, we want to highlight the fact that only cohorts that used full Sanger sequencing were able to detect the RecNciI recombinant allele so far. This once more emphasizes the accuracy of the PacBio sequencing methods for detecting rare and complex *GBA* variants. Additionally, we confirmed that severe variants showed a higher OR than risk variants, which supports the concept of graded risk for different *GBA* variants in PD_GBA_ carriers. ^20^

The most prevalent *GBA* variant in the Luxembourg Parkinson’s study cohort was p. E365K, and the frequency of this variant was similar to what was described in the Irish^20^, Spanish^19^ and New Zealand^5^ populations. It is interesting to note that homozygous carriers of the p.E326K variant do not develop GD.^21^ The variant is associated with PD, and multiple studies have found enrichments varying from 1.60 to 3.34.^22–24^Furthermore, carriers of the risk variants p.E365K and p.T408M were associated with atypical parkinsonism, as these variants were the only ones also present in patients with DLB and PSP in our cohort. Whether this is simply related to the higher frequency of these risk variants in the general population or does have a specific impact on the phenotype needs to be determined in larger studies focusing on *GBA* variants in atypical parkinsonism^25^.

We present a concept for classifying VUS in the *GBA* gene according to the localisation in relation to known variants in sequence and 3D structure, which may help to provide access to future targeted therapies for these patients. Here additional *in vitro* and *ex vivo* studies are needed to functionally validate the impact of these VUS on GCase function in neurons derived from stem cells or in enzyme-activity assays in CSF of affected carriers of these VUS. Additionally, we observed that the average AAO in PD was about four years younger in severe *GBA* carriers compared to non-GBA carriers. This was also observed in previous studies, which showed that PD_GBA_ patients generally have an earlier AAO compared to non-carriers with a median onset in the early fifties.^26,27^

Recent studies have shown that PD_GBA_ carriers have a higher prevalence of cognitive impairment^18,28,29^ and non-motor symptoms including neuropsychiatric disturbances^18,19^, autonomic dysfunction^28^, and sleep disturbances such as RBD^30^. Although not significant after *p*-value adjustment, we found a similar trend and noticed that motor symptoms such as gait disorder, non-motor such as depression and hallucinations symptoms were associated with a more aggressive clinical phenotype in severe *GBA* carriers, supporting the effect of differential *GBA* variant severity.^19,31^

In conclusion, this study showed the utility of targeted PacBio sequencing to identify known and novel *GBA* variants with high accuracy. These findings offer important access to variant-specific counselling. Furthermore, our study describes the full landscape of *GBA* related PD in the current Luxembourgish population showing the high prevalence of *GBA* variants as the major genetic risk in PD.

## 4. Methods

### 4.1. Clinical Cohort

At the time of analysis, the Luxembourg Parkinson’s study comprised 1558 participants (752 patients of parkinsonism and 806 healthy controls (HC) in the frame of the National Centre for Excellence in Research on Parkinson’s disease program (NCER-PD).

All patients complied with the diagnostic criteria of typical PD or atypical parkinsonism as assessed by neurological examination following the United Kingdom Parkinson’s Disease Society Brain Bank (UKPDSBB) diagnostic criteria^32^: 652 fulfilled the criteria for PD, 60 for progressive supranuclear palsy (PSP) including corticobasal syndrome as a subtype of PSP (PSP-CBS), 25 for Dementia with Lewy Body (DLB), 14 for Multiple System Atrophy (MSA), and one for Fronto-temporal dementia with parkinsonism (FTDP). All patients and HC underwent a comprehensive clinical assessment of motor and non-motor symptoms, neuropsychological profile and medical history along with comorbidities. The clinical symptoms assessed, and scales applied are defined in the Supplemental Information^33^. All individuals provided written informed consent. The patients were reassessed at regular follow-up visits. We considered early-onset PD patients those with age at onset (AAO) equal to or younger than 45 years^34^. The genotype-phenotype analysis was based on the assessment of the first visit. The study has been approved by the National Research Ethics Committee (CNER Ref: 201407/13).

### 4.2. Genetic analysis

#### 4.2.1. NeuroChip array

Genotyping was carried out on the InfiniumR NeuroChip Consortium Array^9^ (v.1.0 and v1.1; Illumina, San Diego, CA USA). For rare variants analysis, standard quality control (QC) procedures were conducted, using PLINK v1.9^35^, to remove variants if they had a low genotyping rate (*<*95%) and Hardy-Weinberg equilibrium *p*-value *<* 1×10^−6^. As an additional quality filter, we applied a study-wide allele frequency threshold of <1% in our cohort for rare variants.

#### 4.2.2. GBA-targeted PacBio long-read amplicon sequencing

The targeted *GBA* gene screening was performed by single-molecule real-time (SMRT) long read sequencing^8^ using Sequel II instrument (PacBio). The targeted *GBA* gene coordinates were chr1:155,232,501-155,241,415 (USCS GRCh38/hg38). Long-distance PCR was performed using GBA-specific primer sequences (Forward: 5’-GCTCCTAAAGTTGTCACCCATACATG-3’ and Reverse: 5’-CCAACCTTTCTTCCTTCTTCTCAA-3’)^36^ and the 2x KAPA HiFi Hot Start ReadyMix (Roche). For sample multiplexing, dual asymmetric barcoding was used based on a different 16-bp long index sequence upstream of each of the reverse and forward primers to allow the generation of uniquely barcoded amplicons in one-step PCR amplification. QC was performed prior to pooling. Pools of amplicons were purified with AMPure PacBio beads. A total of 1700 ng of purified amplicon pool was used as input for the SMRTbell library using the SMRTbell Express Template Prep Kit 2.0 (PacBio). Binding of the polymerase and diffusion loading on SMRTCell 8M was prepared according to SMRTLink instructions with CCS reads as sequencing mode (version SMRT Link: 9.0.0.92188). We generated high-quality consensus reads using the PacBio Sequel II sequencer on Circular Consensus Sequencing mode using the pbccs (v6.0.0) tool. The methods replicates both strands of the target DNA.^37^ We demultiplexed and mapped reads from each sample to the human reference genome GRCh38 using minimap2^38^ from the pbmm2 package (v1.4.0) (https://github.com/PacificBiosciences/pbmm2). For variant calling, we used the DeepVariant^39^ (1.0) with models optimized for CCS reads. Finally, we selected variants with quality above 30 (QUAL>30).

#### 4.2.3. Whole genome sequencing

The TruSeq Nano DNA Library Prep Kit (Illumina, San Diego, CA, USA) and MGIEasy FS DNA Prep kit (BGI, China) were used according to the manufacturer’s instructions to construct the WGS library. Paired-end sequencing was performed with the Illumina NovaSeq 6000^40^ and on the MGI G400 sequencers. A QC of the raw data was performed using FastQC (version 0.11.9).^41^ To call the variants, we used the Bio-IT Illumina Dynamic Read Analysis for GENomics (DRAGEN) DNA pipeline^42^ v3.8^43^ with standard parameters. To select the high-quality variants, we annotated and selected variants using VariantAnnotator and SelectVariants modules of the Genome Analysis Toolkit (GATK 4)^44^ pipeline and applied the following additional filtering steps: VariantFiltration module for SNVs (QD<2, FS>60, MQ<40, MQRankSum<-12, ReadPosRankSum<-8, DP<10.0, QUAL<30, VQSLOD<0, ABHet>0.75 or <0.25, SOR>3 and LOD<0), and insertions-deletions (QD<2, FS>200, QUAL<30, ReadPosRankSum<-20, DP<10 and GQ_MEAN<20).

### 4.3. Variant annotation and validation

Variant annotation was done with ANNOVAR,^45^ using the Genome Aggregation Database (gnomAD r2.1)^46^, the Human Gene Mutation Database (HGMD)^47^ and ClinVar^48^, and the Combined Annotation Dependent Depletion (CADD)^49^ and REVEL^50^ to score the pathogenicity of missense variants.^51^ For variants in splice sites, we used the ada_score and rf_score from dbscSNV (version 1.1)^52^. Ada_score ≥ 0.6 or rf_score ≥ 0.6 indicate that the variant is likely to affect splicing.

Rare variants were selected according to minor allele frequency *<* 1% in gnomAD for the Non-Finnish European (NFE) population in the ‘non-neuro’ gnomAD subset. Then, exonic and splicing variants (+/- 2bp from the exon boundary) were selected for autosomal dominant (*LRRK2, SNCA, VPS35, GBA*) and autosomal recessive (*PRKN, PINK1, PARK7, ATP13A2*) PD genes. Rare variants within these genes were then confirmed by Sanger sequencing.^53^

### 4.4. *GBA* variant nomenclature

All variants in *GBA* were annotated based on GRCh37 and were numbered according to the current variant nomenclature guidelines (http://varnomen.hgvs.org), based on the primary translation product (NM_001005742), which includes the 39-residue signal peptide.

### 4.5. *GBA* variant classification

*GBA* variants classification was done according to the PD literature based on the work of Höglinger and colleagues in 2022.^10^ Exonic or splice-site variants that are not mentioned in the paper were subclassified as ‘severe’ *GBA* variants if there were annotated as pathogenic in ClinVar, otherwise they were subclassified as variant with unknown significance (VUS). ^55^

### 4.6 Statistical analysis

To assess the frequency of different *GBA* variant types and analyse the genotype-phenotype associations in the Luxembourg Parkinson’s Study, we considered only unrelated individuals and retained only one proband per family. For cases, we kept the patient with the earliest AAO. We excluded HC with first-degree PD relatives (parents, sibs, and offspring) and age at assessment (AAA) less than 60 years, to account for age-dependent penetrance. Therewith, reduce the gap of age between cases and HC. Thus, 1420 unrelated individuals (742 patients and 678 HC) were selected for the statistical analysis.

We used regression models to assess the effect of PD_GBA_ carrier status on the clinical variables. We excluded individuals carrying only VUS or synonymous variants. To this aim, we performed three types of association tests: (1) all PD_GBA_ pathogenic variant carriers (severe, mild and risk) *vs* PD_GBA-non-carriers_, (2) for each sub-group of PD_GBA_ pathogenic variant carriers *vs* PD_GBA-non-carriers_, (3) severe PD_GBA_ pathogenic variant carriers *vs* combined mild and risk PD_GBA_ pathogenic variant carriers. The effect of each factor was expressed as the Beta (β) regression coefficient. The odds ratio (OR) along with a 95% confidence interval (CI) was used to assess whether a particular exposure is a risk factor for a particular outcome. Regression models were adjusted for AAA, sex, and disease duration. FDR adjusted p-value < 0.05 was considered as statistically significant.

### 4.7 Structure-based evaluation of VUS

To evaluate VUS variants, we implemented a method to assign the pathogenicity based on the REVEL and CADD scores for missense variants and the dbscSNV scores (ada_score and rf_score) for splice variants according to dbNFSP definition^54^, as well as whether the patients carrying the variants.We reclassified a VUS (1) as ‘severe’ if the variant was present only in patients and with deleterious effect in all scores or present only in patients with early onset PD, as ‘mild’ if the variant was present only in patients and with tolerated effect in all scores, as ‘ risk’ if present in patients and HCs or with tolerated and deleterious effect in either score or annotated as ‘PD susceptibility’ in HGMD, and (4) as ‘benign’ if present only in HC. We mapped the known pathogenic missense variants and newly identified VUS identified in our cohort together with all reported population variants from gnomAD onto the *GBA* protein sequence and the 3D structure. We used an X-ray structure of GCase at 2.0 Å resolution (PDB structure accession code 1ogs; https://www.rcsb.org/) (Supplementary Figure 2). The analysis of the 3D structure was carried out by PyMOL (http://www.pymol.org). VUS were evaluated as risk variant if they were 2bp positions away in sequence or had a C-alpha distance of less than 5 ångström in 3D from another known pathogenic variant similar to the approach used in Johannesen et al.^55^

## Data Availability

The dataset for this manuscript is not publicly available as it is linked to the Luxembourg Parkinson's Study and its internal regulations. Any requests for accessing the dataset can be directed to

## 7. Acknowledgments

We would like to thank all participants of the Luxembourg Parkinson’s Study for their important support of our research. Furthermore, we acknowledge the joint effort of the National Centre of Excellence in Research on Parkinson’s Disease (NCER-PD) Consortium members from the partner institutions Luxembourg Centre for Systems Biomedicine, Luxembourg Institute of Health, Centre Hospitalier de Luxembourg, and Laboratoire National de Santé generally contributing to the Luxembourg Parkinson’s Study as listed below. The work presented here was funded by the Luxembourg National Research (FNR/NCER13/BM/11264123), the PEARL program (FNR/P13/6682797 to RK), MotaSYN (12719684 to RK), MAMaSyn (to RK), MiRisk-PD (C17/BM/11676395 to RK, PM), the FNR/DFG Core INTER (ProtectMove, FNR11250962 to PM), and the PARK-QC DTU (PRIDE17/12244779/PARK-QC to RK, SP).

## ON BEHALF OF THE NCER-PD CONSORTIUM

Geeta ACHARYA ^2^, Gloria AGUAYO ^2^, Myriam ALEXANDRE ^2^, Muhammad ALI ^1^, Wim AMMERLANN ^2^, Giuseppe ARENA ^1^, Rudi BALLING ^1^, Michele BASSIS ^1^, Katy BEAUMONT ^2^, Regina BECKER ^1^, Camille BELLORA ^2^, Guy BERCHEM ^3^, Daniela BERG ^11^, Alexandre BISDORFF 5, Ibrahim BOUSSAAD ^1^, Kathrin BROCKMANN ^11^, Jessica CALMES ^2^, Lorieza CASTILLO ^2^, Gessica CONTESOTTO ^2^, Nico DIEDERICH ^3^, Rene DONDELINGER ^5^, Daniela ESTEVES ^2^, Guy FAGHERAZZI ^2^, Jean-Yves FERRAND ^2^, Manon GANTENBEIN ^2^, Thomas GASSER ^11^, Piotr GAWRON ^1^, Soumyabrata GHOSH ^1^, Marijus GIRAITIS ^2,3^, Enrico GLAAB ^1^, Elisa GÓMEZ DE LOPE ^1^, Jérôme GRAAS ^2^, Mariella GRAZIANO ^17^, Valentin GROUES ^1^, Anne GRÜNEWALD ^1^, Wei GU ^1^, Gaël HAMMOT ^2^, Anne-Marie HANFF ^2^, Linda HANSEN ^1^,3, Maxime HANSEN ^1^,^3^, Michael HENEKA ^1^, Estelle HENRY ^2^, Sylvia HERBRINK ^6^, Sascha HERZINGER ^1^, Michael HEYMANN ^2^, Michele HU ^8^, Alexander HUNDT ^2^, Nadine JACOBY ^18^, Jacek JAROSLAW LEBIODA ^1^, Yohan JAROZ ^1^, Quentin KLOPFENSTEIN ^1^, Jochen KLUCKEN ^1, 2^,3, Rejko KRÜGER ^1, 2,3^, Pauline LAMBERT ^2^, Zied LANDOULSI ^1^, Roseline LENTZ ^7^, Inga LIEPELT ^11^, Robert LISZKA ^14^, Laura LONGHINO ^3^, Victoria LORENTZ ^2^, Paula Cristina LUPU ^2^, Clare MACKAY ^10^, Walter MAETZLER ^15^, Katrin MARCUS ^13^, Guilherme MARQUES ^2^, Tainá M MARQUES ^1^, Patricia MARTINS CONDE ^1^, Patrick MAY ^1^, Deborah MCINTYRE ^2^, Chouaib MEDIOUNI ^2^, Francoise MEISCH ^1^, Myriam MENSTER ^2^, Maura MINELLI ^2^, Michel MITTELBRONN ^1,4^, Brit MOLLENHAUER ^12^, Friedrich MÜHLSCHLEGEL ^4^, Romain NATI ^3^, Ulf NEHRBASS ^2^, Sarah NICKELS ^1^, Beatrice NICOLAI ^3^, Jean-Paul NICOLAY ^19^, Marek OSTASZEWSKI ^1^, Clarissa P. da C. GOMES ^1^, Sinthuja PACHCHEK ^1^, Claire PAULY ^1,3^, Laure PAULY ^1^, Lukas PAVELKA ^1,3^, Magali PERQUIN ^2^, Rosalina RAMOS LIMA ^2^, Armin RAUSCHENBERGER ^1^, Rajesh RAWAL ^1^, Dheeraj REDDY BOBBILI ^1^, Kirsten ROOMP ^1^, Eduardo ROSALES ^2^, Isabel ROSETY ^1^, Estelle SANDT ^2^, Stefano SAPIENZA ^1^, Venkata SATAGOPAM ^1^, Margaux SCHMITT ^2^, Sabine SCHMITZ ^1^, Reinhard SCHNEIDER ^1^, Jens SCHWAMBORN ^1^, Jean-Edouard SCHWEITZER ^1^, Amir SHARIFY ^2^, Ekaterina SOBOLEVA ^1^, Kate SOKOLOWSKA ^2^, Olivier TERWINDT ^1^,3, Hermann THIEN ^2^, Elodie THIRY ^3^, Rebecca TING JIIN LOO ^1^, Joana TORRE ^2^, Christophe TREFOIS ^1^, Johanna TROUET ^2^, Olena TSURKALENKO ^2^, Michel VAILLANT ^2^, Mesele VALENTI ^2^, Carlos VEGA ^1^, Liliana VILAS BOAS ^3^, Maharshi VYAS ^1^, Richard WADE-MARTINS ^1^, Paul WILMES ^1^, Evi WOLLSCHEID-LENGELING ^1^, Gelani ZELIMKHANOV ^3^

^1^Luxembourg Centre for Systems Biomedicine, University of Luxembourg, Esch-sur-Alzette, Luxembourg

^2^Luxembourg Institute of Health, Strassen, Luxembourg

3 Centre Hospitalier de Luxembourg, Strassen, Luxembourg

4 Laboratoire National de Santé, Dudelange, Luxembourg

5 Centre Hospitalier Emile Mayrisch, Esch-sur-Alzette, Luxembourg

6 Centre Hospitalier du Nord, Ettelbrück, Luxembourg

7 Parkinson Luxembourg Association, Leudelange, Luxembourg

8 Oxford Parkinson’s Disease Centre, Nuffield Department of Clinical Neurosciences, University of Oxford, Oxford, UK

9 Oxford Parkinson’s Disease Centre, Department of Physiology, Anatomy and Genetics, University of Oxford, Oxford, UK

10 Oxford Centre for Human Brain Activity, Wellcome Centre for Integrative Neuroimaging, Department of Psychiatry, University of Oxford, Oxford, UK

11 Center of Neurology and Hertie Institute for Clinical Brain Research, Department of Neurodegenerative Diseases, University Hospital Tübingen, Tübingen, Germany

12 Paracelsus-Elena-Klinik, Kassel, Germany

13 Ruhr-University of Bochum, Bochum, Germany

14 Westpfalz-Klinikum GmbH, Kaiserslautern, Germany

15 Department of Neurology, University Medical Center Schleswig-Holstein, Kiel, Germany

16 Department of Neurology Philipps, University Marburg, Marburg, Germany

17 Association of Physiotherapists in Parkinson’s Disease Europe, Esch-sur-Alzette, Luxembourg

18 Private practice, Ettelbruck, Luxembourg

19 Private practice, Luxembourg, Luxembourg

## 8. Author contribution

1. Research project: A. Conception, B. Organization, C. Execution;
2. Statistical Analysis: A. Design, B. Execution, C. Review and Critique;
3. Manuscript Preparation: A. Writing of the first draft, B. Review and Critique;
4. Genetic data: A. Sequencing Execution, B. Analysis;
5. Data collection: A. Participation, B. Exportation, C. Curation

SP: 1A, 1B, 1C, 2A, 2B, 2C, 3A, 3B, 4B,5B, 5C

ZL: 1C, 2A, 2B, 2C,3A, 3B, 5C

LP: 1C, 2C, 3B, 5A, 5B, 5C

CS: 2C, 3B, 4A

EBA: 2C, 3B, 4A

CG: 2C, 3B, 4A

AKH: 2C, 3B, 4A

DRB: 2C, 3B

NC: 3B, 4A

PM: 1A, 1B, 1C, 2A, 2C, 3B, 4B, 5C

RK: 1A, 1B, 1C, 2A, 2C, 3B, 5A, 5C

