## Supplementary material for "Accurate long-read sequencing identified GBA variants as a major genetic risk factor in the Luxembourg Parkinson’s study"

**Supplementary. Clinical symptoms and scales**

Clinical symptoms and scales Movement Disorder Society-Unified Parkinson's disease Rating Scale (MDS-UPDRS I-IV) and Scales for Outcomes in Parkinson's Disease-Autonomic questionnaire (SCOPA-AUT) are used under the license number (14017_ND). Probable REM sleep behaviour disorder (pRBD) was based on the validated selfreporting questionnaire REM Sleep Behaviour Disorder Screening Questionnaire (RBDSQ) where possible pRBD was defined as RBDSQ ≥ 7[1]. Assessment of sleep quality was done via Parkinson's Disease Sleep Scale (PDSS)[2]. The calculation of levodopa equivalent daily dose (LEDD, reported in g/day) was based on established conversion factors[3]. Definition of constipation corresponds to the diagnostic criteria ROME III and information was acquired in a semi-structured interview[4]. The Hoehn and Yahr scale (H&Y) corresponds to the modified version of the scale[5]. Quality of life was assessed via Parkinson's disease Questionnaire 39 (PDQ-39)[6]. Depression symptoms were reflected by Beck Depression Inventory Version I (BDI)[7]. Olfactory function was examined with 16 items Sniffin' Stick test[8]. Cognitive performance was assessed via Montreal Cognitive Assessment (MoCA)[9]. Presence of recurrent orthostatic hypotension was assessed using a semi-structured interview inquiring for the symptoms of orthostatic hypotension, i.e. faintness, dizziness, light-headedness, vertigo, hearing disturbance, visual disturbance or syncope following the tilting, standing up or after a long standing relieved by sitting down or laying down. Symptoms included in the analysis (gait disorder, falls, freezing of gait (FOG), dyskinesia, motor fluctuations, excessive daily sleepiness, insomnia, dysphagia, urinary incontinence (corresponding to any type of urinary incontinence, i.e. stress, urge, overflow or mixed urinary incontinence), hallucinations and impulse control disorder (ICD)) were assessed during a semi-structured interview of the participant and/or the participant's proxy with a study physician and refer to the current motor and non-motor symptoms at the time of assessment. DBS, Presence of treatment by Deep Brain Stimulation. Restless leg syndrome corresponds to history or presence of rest less leg syndrome based on the accurate anamnestic description corresponding to the diagnostic features/criteria.

**Supplementary Table 1. Comparison between AAO, AAA and Gender for the Luxembourgish cohort before and after the exclusion criteria.**

| | Before | | After | |
|---|---|---|---|---|
| **Features** | **Cases (n = 752)** | **Controls (n = 806)** | **Cases (n = 742)** | **Controls (n = 678)** |
| **AAA** | 67.7 (±10.5) | 59.3 (±12.2) | 67.8 (±10.5) | 61 (±11.5) |
| **AAO, mean (SD)** | 63.1 (±16) | - | 63.1 (±11.5) | - |
| **Sex, Male % (n)** | 499 (66.4%) | 426 (52.9%) | 494 (66.6%) | 371 (54.7%) |

We applied a t-test for AAA and AAO features and a Fisher test for the gender features.

**Supplementary Table 2. PCR primers**

PCR Primers (Stone et al. 2000)

| exon | Forward | reverse | length |
|---|---|---|---|
| 1–5 | CCTAAAGTTGTCACCCATAC | AGCAGACCTACCCTACAGTTT | 2972 |
| 5–7 | GACCTCAAATGATATACCTG | AGTTTGGGAGCCAGTCATTT | 2049 |
| 8–11 | TGTGTGCAAGGTCCAGGATCAG | ACCACCTAGAGGGGAAAGTG | 1682 |

Sequencing Primers (partly Stone et al.2000)

| exon | Forward | Reverse | Length |
|---|---|---|---|
| 1 | CCTAAAGTTGTCACCCATAC | aaattccagtgccaggattc | 392 |
| 2 | GAGAGTAGTTGAGGGGTGGA | CAAAGGACTATGAGGCAGAA | 210 |
| 3 | ATGTGTCCATTCTCCATGTC | GGTGATCACTGACACCATTT | 323 |
| 4 | GGTGTCAGTGATCACCATGG | ACGAAAAGTTTCAATGGCTCT | 263 |
| 5 | GCAAGTGATAAGCAGAGTCC | AGCAGACCTACCCTACAGTTT | 280 |
| 6 | CTCTGGGTGCTTCTCTCTTC | ACAGATCAGCATGGCTAAAT | 271 |
| 7 | TTGGCCGGATCATTCATGAC | AGTTTGGGAGCCAGTCATTT | 342 |
| 8 | TGTGTGCAAGGTCCAGGATCAG | TTTGCAGGAAGGGAGACTGG | 294 |
| 9 | CACAGGGCTGACCTACCCAC | GCTCCCTCGTGGTGTAGAGT | 307 |
| 10 | CAGGAGTTATGGGGTGGGTC | GAGGCACATCCTTAGAGGAG | 329 |
| 11 | GTGGGCTGAAGACAGCGTTGG | ACCACCTAGAGGGGAAAGTG | 342 |

The first step is a long-range PCR which ensures that the pseudogene of GBA is <u>not</u> amplified.

**Supplementary Table 3. Exonic and splice-site *GBA* variants found in the Luxembourg Parkinson's study.**

| Subclassification | protein change | Nucleotide change | dbSNP | Variant type | Höglinger et al. | ClinVar Significance | ClinVar | HGMD | REVEL | CADD | gnomAD NFE | 3D | domain |
|---|---|---|---|---|---|---|---|---|---|---|---|---|---|
| **severe** | - | c.115+1G>A | rs104886460 | splicing | severe | Pathogenic | PD/GD/DLB | DM | - | D | rare | / | / |
| | p.P161S | c.C481T | rs121908299 | missense | - | Pathogenic | GD | DM | D | D | - | β-sheet | III |
| | p.G234W | c.G700T | - | missense | severe | - | - | DM | D | D | - | coil-loop | III |
| | p.G241R | c.G721A | rs409652 | missense | severe | Pathogenic | GD | DM | D | T | rare | coil-turn | III |
| | p.F252I | c.T754A | rs381737 | missense | severe | Pathogenic | GD | DM | D | T | rare | α-helix | III |
| | p.H294Q | c.T882G | rs367968666 | missense | severe | Pathogenic | GD | DM | T | T | | α-helix | III |
| | p.R398* | c.C1192T | rs121908309 | nonsense | severe | Pathogenic | GD | DM | - | D | - | α-helix | III |
| | p.G416S | c.G1246A | rs121908311 | missense | severe | Pathogenic | GD | DM | D | T | rare | β-sheet | III |
| | p.L483P | c.T1448C | rs421016 | missense | severe | Pathogenic | GD | DM | D | T | rare | β-sheet | II |
| | p.R502H | c.G1505A | rs80356772 | missense | severe | Conflicting interpretations of pathogenicity | GD | DM | D | D | rare | coil-loop | II |
| **mild** | p.N409S | c.A1226G | rs76763715 | missense | mild | Pathogenic | PD/GD/DLB | DM | T | T | rare | α-helix | III |
| **risk** | p.E365K | c.G1093A | rs2230288 | missense | risk | Benign | GD | DM | T | T | 1.4% | α-helix | III |
| | p.T408M | c.C1223T | rs75548401 | missense | risk | Conflicting interpretations of pathogenicity | PD/GD | DM | T | T | 1.1% | buried residue | III |
| **VUS** | p.K13R | c.A38G | rs150466109 | missense | VUS | Benign | GD | DM | T | T | rare | / | / |
| | p.Y61H | c.T181C | rs1266341749 | missense | - | - | - | - | T | T | rare | coil-loop | I |
| | p.R78C | c.C232T | rs146774384 | missense | - | - | - | DM | T | D | rare | β-sheet | II |
| | p.A97G | c.C290G | - | missense | - | - | - | - | T | T | - | coil-bend | II |
| | p.L213P | c.T638C | - | missense | VUS | - | - | DM | D | D | - | β-sheet | III |
| | p.A215D | c.C644A | - | missense | - | - | - | DM | D | D | - | β-sheet | III |
| | p.E427K | c.G1279A | rs149171124 | missense | VUS | Uncertain significance | Parkinsonism | DM | T | T | rare | coil-turn | I |
| | p.R434C | c.C1300T | rs747284798 | missense | - | - | - | DM | D | D | - | coil-loop | I |
| | p.A495P | c.G1483C | rs368060 | missense | - | Benign | GD | DM | T | T | rare | β-sheet | II |
| | p.H529R | c.A1586G | - | missense | VUS | - | - | DM | T | T | - | β-sheet | II |
| | p.R534C | c.C1600T | rs146519305 | missense | - | - | - | - | T | T | rare | coil-loop | II |
| | p.T408T | c.G1224A | rs138498426 | synonymous | VUS | Uncertain significance | GD | DM | - | - | rare | buried residue | III |

Abbreviations: GBA, glucocerebrosidase gene; GD, Gaucher's disease; PD, Parkinson's disease; DLB, Dementia with Lewy Bodies. HGMD, The Human Gene Mutation Database; REVEL, Rare Exome Variant Ensemble Learner; CADD, Combined Dependent Depletion; gnomAD, The Genome Aggregation Database. DM, Disease causing mutation; D, Deleterious; T, Tolerate; VUS, Variants of unknown significant.

**Supplementary Table 4. Exonic synonymous GBA variants in the Luxembourg Parkinson's study.**

| Subjects | | nucleotide change | protein change | dbSNP | Exon | 3D | gnomAD NFE | ClinVar | ClinVar Significance | HGMD |
|---|---|---|---|---|---|---|---|---|---|---|
| PD | HC | | | | | | | | | |
| 5 | 3 | c.G1497C | p.V499V | rs1135675 | 11 | β-sheet | rare | - | - | - |
| | 1 | c.C1473A | p.P491P | rs149257166 | 11 | coil-turn | - | - | - | - |
| 1 | | c.A1455G | p.A485A | rs199928507 | 11 | β-sheet | rare | - | - | - |
| 1 | | c.C228T | p.F76F | rs75954905 | 4 | β-sheet | rare | - | - | - |
| 2 | 1 | c.G1224A | p.T408T | rs138498426 | 9 | buried residue | rare | GD | Uncertain significance | PD |
| 1 | | c.T1029C | p.Y343Y | - | 9 | coil-turn | - | - | - | - |
| 2 | | c.C630T | p.P210P | rs201615998 | 7 | coil-loop | rare | - | - | - |
| 1 | | c.C585G | p.L195L | rs1157873928 | 6 | π-helix | - | - | - | - |
| 1 | | c.G105A | p.S35S | rs148001886 | 3 | - | rare | - | - | - |

All variants were identified in the heterozygous state.
Abbreviations: GBA, glucocerebrosidase gene; GD, Gaucher's disease; PD, Parkinson's disease; HC, Healthy controls; HGMD, The Human Gene Mutation Database ; gnomAD, The Genome Aggregation Database.

**Supplementary Table 5. Splicing, intronic and UTRs regions variants detected in the Luxembourg Parkinson’s study by PacBio sequencing method.**

| N° | POS | REF | ALT | Region | Transcript | Nucleotide changes | GnomAD NFE | avsnp150 | ClinVar | ClinVar Significance | PD | HC |
|---|---|---|---|---|---|---|---|---|---|---|---|---|
| 1 | 155204345 | C | T | UTR3 | NM_001005742 | c.*441G>A | | | | | 1 | 0 |
| 2 | 155204541 | G | A | UTR3 | NM_001005743 | c.*245C>T | 0.0002 | | | | 0 | 1 |
| 3 | 155204621 | A | G | UTR3 | NM_001005744 | c.*165T>C | 0.0393 | rs375776699 | | | 81 | 79 |
| 4 | 155204684 | A | G | UTR3 | NM_001005745 | c.*102T>C | 0.0007 | rs368275143 | | | 17 | 21 |
| 5 | 155204694 | C | T | UTR3 | NM_001005746 | c.*92G>A | 0.0025 | rs708606 | GD/DLB | Likely_benign | 18 | 23 |
| 6 | 155204701 | C | T | UTR3 | NM_001005747 | c.*85G>A | | | | | 0 | 1 |
| 7 | 155205200 | G | T | intronic | | | 0.0018 | rs183510604 | | | 0 | 2 |
| 8 | 155205203 | G | A | intronic | | | 0.0003 | rs426516 | | | 4 | 1 |
| 9 | 155205300 | G | A | intronic | | | | | | | 3 | 1 |
| 10 | 155205359 | G | A | intronic | | | | | | | 0 | 1 |
| 11 | 155205378 | C | T | intronic | | | 0.0139 | rs12752133 | | | 30 | 16 |
| 12 | 155205646 | G | C | intronic | | | 6.668e-05 | | | | 1 | 0 |
| 13 | 155205669 | G | T | intronic | | | 0.9992 | rs3115534 | GD | Benign | 752 | 806 |
| 14 | 155205709 | A | G | intronic | | | 0.0007 | rs548435731 | | | 0 | 1 |
| 15 | 155205748 | G | A | intronic | | | 6.681e-05 | rs1003268223 | | | 1 | 1 |
| 16 | 155205801 | A | G | intronic | | | | | | | 1 | 0 |
| 17 | 155205964 | C | T | intronic | | | | | | | 0 | 1 |
| 18 | 155206430 | G | A | intronic | | | 6.688e-05 | rs767239225 | | | 0 | 1 |
| 19 | 155206542 | TTGTGTGTGTA | T | intronic | | | | rs998227221 | | | 0 | 1 |
| 20 | 155206578 | G | GTA | intronic | | | 0.0003 | rs200655080 | | | 1 | 3 |
| 21 | 155206580 | A | G | intronic | | | 6.711e-05 | rs146697312 | | | 1 | 0 |
| 22 | 155206863 | C | G | intronic | | | 6.704e-05 | rs1026559493 | | | 1 | 0 |
| 23 | 155206981 | C | T | intronic | | | | | | | 1 | 0 |
| 24 | 155207030 | G | A | intronic | | | 0.0233 | rs72704130 | | | 36 | 52 |
| 25 | 155207050 | G | A | intronic | | | | rs749925127 | | | 1 | 0 |
| 26 | 155207387 | A | T | intronic | | | 0.015 | rs140335079 | PD/GD/DLB | Benign | 14 | 27 |
| 27 | 155207449 | C | T | intronic | | | | rs1006437355 | | | 2 | 0 |
| 28 | 155207550 | G | T | intronic | | | 0.0001 | | | | 0 | 2 |
| 29 | 155207674 | C | CAG | intronic | | | | | | | 1 | 0 |
| 30 | 155207733 | C | T | intronic | | | 0.0396 | rs28678003 | | | 82 | 79 |
| 31 | 155207846 | T | C | intronic | | | 0.0002 | rs145066479 | | | 3 | 1 |
| 32 | 155207848 | G | T | intronic | | | 0.0043 | rs183540501 | | | 4 | 2 |
| 33 | 155207866 | A | T | intronic | | | | rs529870563 | | | 1 | 2 |
| 34 | 155208167 | G | A | intronic | | | | rs566671462 | | | 0 | 2 |
| 35 | 155208495 | G | A | intronic | | | 0.0003 | rs567935648 | | | 0 | 1 |
| 36 | 155208519 | CT | C | intronic | | | 6.673e-05 | | | | 1 | 2 |
| 37 | 155208611 | A | G | intronic | | | 0.0003 | rs569282073 | | | 2 | 1 |
| 38 | 155208624 | A | G | intronic | | | 0.0129 | rs188328778 | | | 15 | 17 |
| 39 | 155208644 | C | T | intronic | | | 6.675e-05 | | | | 0 | 1 |
| 40 | 155208647 | T | C | intronic | | | 0.7089 | rs7416991 | | | 687 | 734 |
| 41 | 155208647 | T | G | intronic | | | 0.2911 | rs7416991 | | | 316 | 349 |
| 42 | 155208647 | T | G | intronic | | | 0.2911 | rs7416991 | | | 65 | 72 |
| 43 | 155208805 | C | T | intronic | | | 0.0 | | | | 1 | 0 |
| 44 | 155208851 | T | C | intronic | | | 6.774e-05 | rs149120852 | | | 0 | 1 |
| 45 | 155209078 | C | T | intronic | | | 0.0 | rs1005434278 | | | 1 | 0 |
| 46 | 155209079 | G | A | intronic | | | 0.0001 | rs114452199 | | | 3 | 2 |
| 47 | 155209082 | A | G | intronic | | | | rs899199374 | | | 1 | 2 |
| 48 | 155209251 | G | A | intronic | | | | rs991547343 | | | 1 | 0 |
| 49 | 155209297 | C | T | intronic | | | 0.0011 | rs183903019 | | | 4 | 3 |
| 50 | 155209298 | G | A | intronic | | | 0.0002 | rs776425625 | | | 0 | 2 |
| 51 | 155209594 | G | A | intronic | | | 0.0001 | rs377315750 | | | 1 | 0 |
| 52 | 155209913 | T | G | intronic | | | 0.0005 | rs199565854 | | | 0 | 2 |
| 53 | 155209938 | G | A | intronic | | | 0.0003 | rs559516544 | | | 5 | 5 |
| 54 | 155209962 | C | T | intronic | | | 0.0115 | rs114217696 | | | 14 | 12 |
| 55 | 155210030 | G | A | intronic | | | 0.0001 | rs151028758 | | | 3 | 2 |
| 56 | 155210070 | C | CA | intronic | | | | | | | 0 | 1 |
| 57 | 155210146 | C | T | intronic | | | 0.0001 | | | | 2 | 1 |
| 58 | 155210156 | G | C | intronic | | | | | | | 0 | 1 |
| 59 | 155210170 | G | A | intronic | | | 0.0002 | rs962460364 | | | 0 | 1 |
| 60 | 155210570 | T | C | intronic | | | 0.001 | rs2361534 | | | 0 | 2 |
| 61 | 155210613 | C | T | intronic | | | | | | | 1 | 1 |
| 62 | 155210641 | A | C | intronic | | | 0.0002 | rs2070679 | | | 3 | 3 |
| 63 | 155210723 | C | T | intronic | | | | | | | 1 | 0 |
| 64 | 155210739 | C | T | intronic | | | | | | | 1 | 0 |
| 65 | 155210918 | T | C | UTR5 | NM_001005742 | c.-15A>G | 0.0013 | rs41264927 | | | 3 | 2 |
| 66 | 155211027 | C | T | UTR5 | NM_000157 | c.-124G>A | | | | | 0 | 1 |
| 67 | 155211089 | C | T | intronic | | | | | | | 1 | 0 |
| 68 | 155211101 | G | A | intronic | | | 0.0 | rs1007847984 | | | 0 | 1 |
| 69 | 155211106 | T | C | intronic | | | 0.0116 | rs188978150 | | Uncertain_significance | 30 | 27 |

Abbreviations: GD, Gaucher’s disease; PD, Parkinson’s disease; DLB, Dementia with Lewy Bodies.; gnomAD, The Genome Aggregation Database. DM, Disease causing mutation; FP, in vitro or in vivo functional polymorphism.

### Supplementary Table 6.

| Type of data | Clinical characteristics and scales | PD GBA carrier | | missing values (%) | β (95%) | *p*-value | adj *p*-value |
|---|---|---|---|---|---|---|---|
| | | **Yes (n=67)** | **No (N=561)** | | | | |
| Motor symptoms/scales | H&Y, mean (SD) | 2.2 (±0.8) | 2.2 (±0.8) | 3 (0.5%) | 0.1 (-0.08 to 0.27) | 0.2775 | 0.9281 |
| | MDS-UPDRS II, mean (SD) | 11.4 (±7.3) | 11.4 (±8.3) | 13 (2.1%) | 0.29 (-1.58 to 2.16) | 0.7596 | 0.9603 |
| | MDS-UPDRS III, mean (SD) | 33.1 (±16.5) | 34.6 (±16.2) | 15 (2.4%) | -0.81 (-4.73 to 3.11) | 0.6849 | 0.9603 |
| | MDS-UPDRS IV, mean (SD) | 1.6 (±3.2) | 1.7 (±3.4) | 7 (1.1%) | -0.05 (-0.81 to 0.71) | 0.8904 | 0.9603 |
| | Dyskinesias, n (%) | 9 (13.4%) | 67 (11.9%) | 0 | 0.2 (-0.62 to 1.01) | 0.6358 | 0.9603 |
| | Falls, n (%) | 12 (17.9%) | 98 (17.5%) | 0 | 0.13 (-0.57 to 0.83) | 0.7182 | 0.9603 |
| | Gait Disorder, n (%) | 37 (55.2%) | 314 (56.0%) | 0 | 0.03 (-0.49 to 0.56) | 0.8982 | 0.9603 |
| | FOG, n (%) | 15 (22.4%) | 126 (22.5%) | 0 | 0.06 (-0.61 to 0.74) | 0.8562 | 0.9603 |
| | Restless leg syndrome, n (%) | 10 (14.9%) | 46 (8.2%) | 0 | 0.65 (-0.1 to 1.39) | 0.0874 | 0.9281 |
| | Motor fluctuation, n (%) | 10 (14.9%) | 95 (16.9%) | 0 | -0.17 (-0.94 to 0.61) | 0.674 | 0.9603 |
| Non-motor symptoms/scales | BDI, mean (SD) | 9.4 (±5.8) | 9.9 (±7.1) | 30 (4.8%) | -0.56 (-2.32 to 1.19) | 0.5286 | 0.9603 |
| | MDS-UPDRS Part I, mean (SD) | 11.1 (±6.8) | 10.6 (±7.0) | 15 (2.4%) | 0.59 (-1.11 to 2.29) | 0.4965 | 0.9603 |
| | PDSS, mean (SD) | 103.0 (±22.8) | 104.7 (±24.9) | 46 (7.3%) | -1.82 (-8.06 to 4.42) | 0.5676 | 0.9603 |
| | SCOPA-AUT, mean (SD) | 14.9 (±7.8) | 15.0 (±8.1) | 34 (5.4%) | 0.08 (-1.89 to 2.06) | 0.9329 | 0.9603 |
| | **Sniffin's stick test, mean (SD)** | **7.0 (±3.7)** | **7.8 (±3.6)** | **8 (1.3%)** | **-1.04 (-1.92 to -0.17)** | **0.0198*** | **0.693** |
| | SAS, mean (SD) | 13.9 (±5.8) | 14.0 (±5.7) | 36 (5.7%) | 0.02 (-1.44 to 1.48) | 0.9801 | 0.9801 |
| | MoCA, mean (SD) | 24.7 (±4.2) | 24.4 (±4.5) | 14 (2.2%) | -0.06 (-1.12 to 0.99) | 0.9048 | 0.9603 |
| | Constipation, n (%) | 29 (43.3%) | 251 (44.7%) | 0 | -0.03 (-0.55 to 0.49) | 0.9198 | 0.9603 |
| | Dysphagia, n (%) | 15 (22.4%) | 146 (26.0%) | 0 | -0.19 (-0.8 to 0.42) | 0.5486 | 0.9603 |
| | Insomnia, n (%) | 17 (25.4%) | 154 (27.5%) | 0 | -0.14 (-0.73 to 0.45) | 0.6443 | 0.9603 |
| | Orthostatism, n (%) | 24 (35.8%) | 166 (29.6%) | 0 | 0.31 (-0.23 to 0.84) | 0.2635 | 0.9281 |
| | Urinary incontinence, n (%) | 25 (37.3%) | 171 (30.5%) | 0 | 0.36 (-0.18 to 0.9) | 0.1874 | 0.9281 |
| | Hallucinations, n (%) | 16 (23.9%) | 87 (15.5%) | 0 | 0.6 (-0.02 to 1.22) | 0.0598 | 0.9281 |
| | Excessive daytime sleepiness, n (%) | 23 (34.3%) | 176 (31.4%) | 0 | 0.18 (-0.36 to 0.73) | 0.5122 | 0.9603 |
| | ICD, n (%) | 6 (9.0%) | 55 (9.8%) | 0 | -0.07 (-0.98 to 0.84) | 0.8819 | 0.9603 |
| | Syncope, n (%) | 6 (9.0%) | 27 (4.8%) | 0 | 0.76 (-0.18 to 1.71) | 0.114 | 0.9281 |
| | RBDSQ, mean (SD) | 25 (37.3%) | 171 (30.5%) | 45 (7.2%) | 0.39 (-0.18 to 0.96) | 0.179 | 0.9281 |
| Other clinical outcome | LEDD (mg/day), mean (SD) | 543.4 (±442.6) | 514.7 (±404.7) | 21 (3.3%) | 36.38 (-53.26 to 126.02) | 0.4263 | 0.9603 |
| | PDQ-39, mean (SD) | 39.5 (±27.3) | 39.6 (±26.8) | 51 (8.1%) | -0.37 (-6.76 to 6.03) | 0.9107 | 0.9603 |
| | DBS, n (%) | 4 (6.0%) | 24 (4.3%) | 0 | 0.68 (-0.58 to 1.94) | 0.2887 | 0.9281 |
| Comorbidities | Diabetes, n (%) | 8 (11.9%) | 55 (9.8%) | 0 | 0.35 (-0.46 to 1.16) | 0.3952 | 0.9603 |
| | Hypercholesterolemia, n (%) | 25 (37.3%) | 228 (40.6%) | 0 | -0.1 (-0.64 to 0.43) | 0.7031 | 0.9603 |
| | Cardiovascular disease, n (%) | 10 (14.9%) | 118 (21.0%) | 0 | -0.3 (-1.03 to 0.42) | 0.4134 | 0.9603 |
| | Arterial hypertension, n (%) | 23 (34.3%) | 250 (44.6%) | 0 | -0.38 (-0.93 to 0.17) | 0.1718 | 0.9281 |
| | Traumatic Brain Injury, n (%) | 11 (16.4%) | 124 (22.1%) | 0 | -0.37 (-1.04 to 0.31) | 0.2917 | 0.9281 |

We consider severe, mild, and risk GBA variants as pathogenic mutations. We used regression models (linear and logistic). Data are given as mean and standard deviation (SD) for continuous clinical outcomes and as percentage for binary clinical outcomes. Models adjusted for sex, age at assessment, and disease duration. Beta (β) regression coefficient are given with the 95% CI. Statistically significant results highlighted in bold with (*) sign and red (p-value < 0.05). Abbreviation : p-value, unadjusted p-value; adj p-value, corrected for multiple comparisons using FDR adjustment; AAO, age at onset; H&Y, Hoehn & Yahr; MDS-UPDRS, Movement Disorders Society - Unified Parkinson's Disease Rating Scale; FOG, freezing of gait; BDI, Beck Depression Inventory; PDSS, Panic Disorder Severity Scale; SCOPA-AUT, Scales for Outcomes in Parkinson's Disease-Autonomic questionnaire; SAS, Starkstein apathy scale; MoCA, Montreal Cognitive Assessment; ICD, impulse control disorder; RBDSQ, REM Sleep Behavior Disorder Screening Questionnaire; LEDD, L-dopa equivalent daily dose (mg/day); PDQ-39, Parkinson's Disease quality of life Questionnaire; DBS, Presence of treatment by Deep Brain Stimulation.

**Supplementary Table 7. Frequency of GBA variant in European Parkinson's disease population that performed full GBA gene sequencing.**

| Population | PD (n) | Screening method | GBA carrier frequency (%) | E365K | T408M | L483P | N409S | RecNcil | Other |
|---|---|---|---|---|---|---|---|---|---|
| **Ashkenazi Jews**[10] | 735 | Targeted NGS | 18 | 1.6 | 0 | 0.3 | 11.8 | 0 | 4.2 |
| **Netherland**[11] | 3402 | Long-range PCR | 15 | 6.7 | 2.5 | 0.6 | 0.9 | 0 | 4.3 |
| **Italy**[12] | 874 | Complete exon Sanger sequencing | 14.3 | 1.7 | 0.6 | 2.3 | 3.3 | 0.8 | 5.3 |
| **Luxembourg (this cohort)** | **644** | **PacBio** | **12** | **3.6** | **2.6** | **1.7** | **1.1** | **0.6** | **3** |
| **Southern Spain**[13] | 532 | High-resolution melting analysis (HRM) | 11.7 | 3 | 0.9 | 2.4 | 0.9 | 0 | 4.3 |
| **New Zealand**[14] | 229 | PCR amplicon + nanopore | 9.2 | 4.8 | 3.1 | 0 | 0.4 | 0 | 1.3 |
| **Ireland**[15] | 314 | Complete exon Sanger sequencing | 8.3 | 4.1 | 1.9 | 0 | 0.9 | 0.9 | 0.5 |
| **Portugal**[16] | 230 | X | 8.3 | 0.9 | 0.9 | 1.3 | 2.2 | 0 | 0.9 |
| **Greece**[17] | 172 | X | 6.4 | 0.6 | 0 | 1.2 | 0 | 0 | 4.6 |

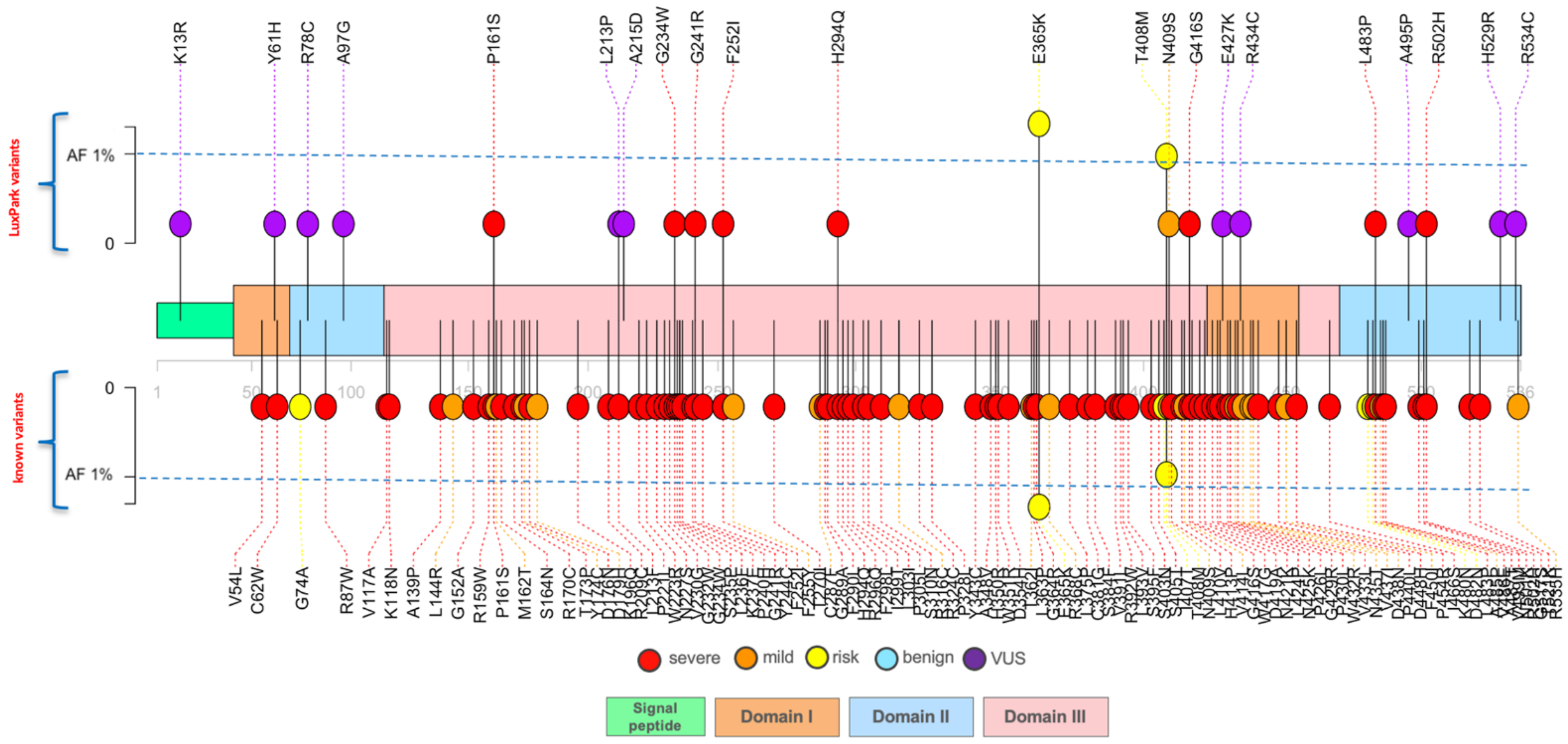


**Supplementary Figure 1:** Lolliplot graphics showing all GBA variants found in the Luxembourg Parkinson's study through the whole protein sequences. Each lollipop represents a GBA variant identified in this the Luxembourgish cohort (upper part) and known GBA variants in ClinVar or literatures (bottom part). Variants classified as severe are colored red, mild are colored orange, risk in yellow and VUS are colored purple.

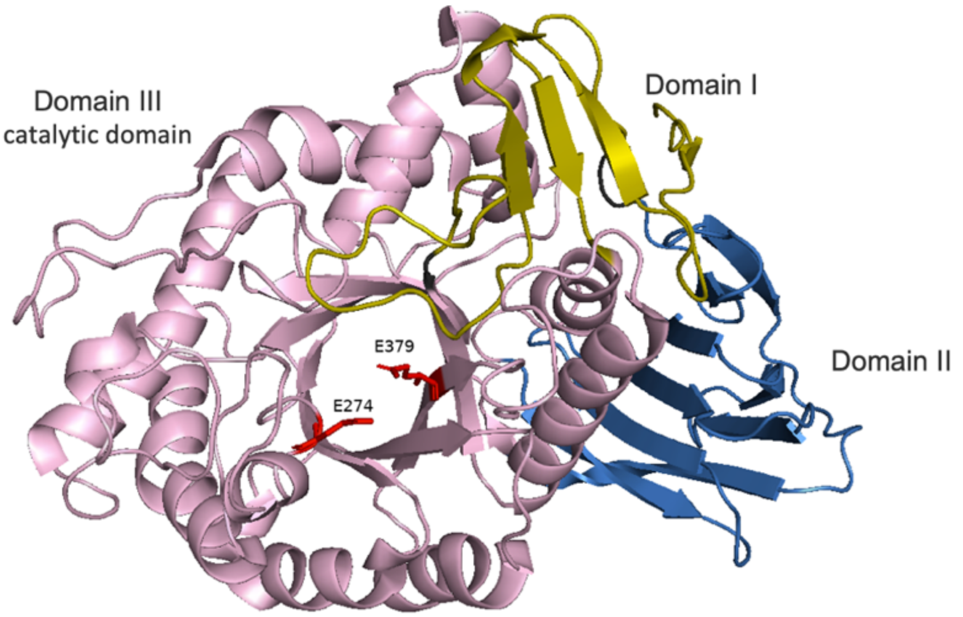


**Supplementary Figure 2**. The 3D structure of GCase (PDB code 1ogs) created using PYMOL (http://www.pymol.org). Domain I is shown in dark yellow with the antiparallel β sheet (residues 1–27 and 383–414), Domain II in blue with the immunoglobulin-like domain (residues 30–75 and 431–497), and Domain III in pink is the catalytic domain with the (b/a)8 (TIM) barrel structure (residues 76–381 and 416–430). The active site residues Glu274 and Glu379 are shown in red.

**References to supplementary material**


1. Nomura T, Inoue Y, Kagimura T, Uemura Y, Nakashima K. Utility of the REM sleep behavior disorder screening questionnaire (RBDSQ) in Parkinson's disease patients. *Sleep Med*. 2011;12(7):711-713. doi:10.1016/j.sleep.2011.01.015
2. Chaudhuri KR, Pal S, Dimarco A, et al. The Parkinson's disease sleep scale: a new instrument for assessing sleep and nocturnal disability in Parkinson's disease. *J Neurol Neurosurg Psychiatry*. 2002;73:629-635. doi:10.1136/jnnp.73.6.629
3. Tomlinson CL, Stowe R, Patel S, Rick C, Gray R, Clarke CE. Systematic review of levodopa dose equivalency reporting in Parkinson's disease. *Movement Disorders*. 2010;25(15):2649-2653. doi:10.1002/mds.23429
4. Longstreth GF, Thompson WG, Chey WD, Houghton LA, Mearin F, Spiller RC. Functional Bowel Disorders. *Gastroenterology*. 2006;130(5):1480-1491. doi:10.1053/j.gastro.2005.11.061
5. Goetz CG, Poewe W, Rascol O, et al. Movement Disorder Society Task Force report on the Hoehn and Yahr staging scale: Status and recommendations. *Movement Disorders*. 2004;19(9):1020-1028. doi:10.1002/mds.20213
6. Peto V, Jenkinson C, Fitzpatrick R, Greenhail R. *The Development and Validation of a Short Measure of Functioning and Well Being for Individuals with Parkinson's Disease*.
7. Beck AT, Ward CH, Mendelson M, Mock J, Erbaugh J. *An Inventory for Measuring Depression The Difficulties Inherent in Obtaining*. http://archpsyc.jamanetwork.com/
8. Hummel T, Kobal G, Gudziol H, Mackay-Sim A. Normative data for the "Sniffin'' Sticks" including tests of odor identification, odor discrimination, and olfactory thresholds: An upgrade based on a group of more than 3,000 subjects." *European Archives of Oto-Rhino-Laryngology*. 2007;264(3):237-243. doi:10.1007/s00405-006-0173-0
9. Nasreddine ZS, Phillips NA, Bédirian V, et al. *The Montreal Cognitive Assessment, MoCA: A Brief Screening Tool For Mild Cognitive Impairment*. www.mocatest.
10. Ruskey JA, Greenbaum L, Roncière L, et al. Increased yield of full GBA sequencing in Ashkenazi Jews with Parkinson's disease. *Eur J Med Genet*. 2019;62(1). doi:10.1016/j.ejmg.2018.05.005
11. den Heijer JM, Cullen VC, Quadri M, et al. A Large-Scale Full GBA1 Gene Screening in Parkinson's Disease in the Netherlands. *Movement Disorders*. 2020;35(9):1667-1674. doi:10.1002/mds.28112
12. Petrucci S, Ginevrino M, Trezzi I, et al. GBA-Related Parkinson's Disease: Dissection of Genotype–Phenotype Correlates in a Large Italian Cohort. *Movement Disorders*. 2020;35(11). doi:10.1002/mds.28195
13. Jesús S, Huertas I, Bernal-Bernal I, et al. GBA variants influence motor and non-motor features of Parkinson's disease. *PLoS One*. 2016;11(12). doi:10.1371/journal.pone.0167749
14. Graham OEE, Pitcher TL, Liau Y, et al. Nanopore sequencing of the glucocerebrosidase (GBA) gene in a New Zealand Parkinson's disease cohort. *Parkinsonism Relat Disord*. 2020;70:36-41. doi:10.1016/j.parkreldis.2019.11.022
15. Olszewska DA, McCarthy A, Soto-Beasley AI, et al. Association Between Glucocerebrosidase Mutations and Parkinson's Disease in Ireland. *Front Neurol*. 2020;11. doi:10.3389/fneur.2020.00527
16. Bras J, Paisan-Ruiz C, Guerreiro R, et al. Complete screening for glucocerebrosidase mutations in Parkinson disease patients from Portugal. *Neurobiol Aging*. 2009;30(9):1515-1517. doi:10.1016/j.neurobiolaging.2007.11.016

17. Kalinderi K, Bostantjopoulou S, Paisan-Ruiz C, Katsarou Z, Hardy J, Fidani L. Complete screening for glucocerebrosidase mutations in Parkinson disease patients from Greece. *Neurosci Lett*. 2009;452(2):87-89. doi:10.1016/j.neulet.2009.01.029
